# Comprehensive neurodevelopmental assessment through non-specialists: Validation of the STREAM digital platform in India and Malawi

**DOI:** 10.64898/2025.12.03.25341202

**Authors:** Elin H Williams, Gareth McCray, Nicholas M Thompson, Maria M Crespo-Llado, Alok Ranjan, Diksha Gajria, Debarati Mukherjee, Vukiwe Ngoma, Chisomo Namathanga, Richard Nkhata, Allan Bennie, Ulemu Kawelama, Naina Midha, Anindita Singh, Innocent Mpakiza, Teresa Del Bianco, Georgia Lockwood Estrin, Luke Mason, Abhishek Chaudhary, Sheffali Gulati, Mark H Johnson, Matthew K Belmonte, Emily Jones, Vikram Patel, Sharat Chandran, Supriya Bhavnani, Emmie Mbale, Gauri Divan, Melissa Gladstone, Bhismadev Chakrabarti

## Abstract

**Background:** Children in low-resource settings often lack access to culturally appropriate and feasible neurodevelopmental assessments. Existing tools are costly, require specialist training, and were developed in high-resource settings, limiting their scalability. The Scalable TRansdiagnostic Early Assessment of Mental health (STREAM) project uses a mobile platform, delivered by non-specialist workers, to assess motor, social, and cognitive development in children aged 0-6 years. We report STREAM scores, psychometric properties, and age-standardised reference curves from India and Malawi.

**Methods:** We recruited children aged 0-6 years in New Delhi (India) and Blantyre (Malawi) from two samples: community (clinics and early education centres) and enriched (children with existing/suspected neurodevelopmental conditions). Trained non-specialists administered STREAM in community settings. Motor, cognitive, and social domain scores were derived from age-adjusted data using structural equation modelling (SEM). Validity was assessed against the Griffiths Mental Development Scales (GMDS), caregiver-reported developmental measures, and anthropometric indices. We examined known-groups discriminability, test-retest reliability, and 18-month responsiveness to change.

**Findings:** We assessed 3992 children (community=3733; enriched=259). SEM fit exceeded standard benchmarks (RMSEA=0.03; CFI=0.98; TLI=0.97). Across both sites, we found extensive evidence for STREAM’s criterion validity [range: *r*=0.31,0.65], convergent validity (9/12 related constructs correlated significantly), and known-groups validity (all hypothesised groups differed significantly). Test-retest reliability was largely good (>0.60&<0.74), and the Indian sample showed some evidence of responsiveness to change.

**Interpretation:** We provide evidence that STREAM is a valid and reliable mobile assessment of neurodevelopment in low-resource settings, designed for delivery by non-specialists in community settings. It captures key neurodevelopmental domains, demonstrates robust psychometric properties, enables standardised monitoring via reference curves, and provides a scalable approach to reducing inequities in access to neurodevelopmental assessments in settings where specialist-delivered tools may not be feasible due to cost, training requirements, or cultural applicability.

**Funding:** Medical Research Council (MRC) Global Challenges Research Fund (MR/S036423/1).

## Introduction

The early years of life form a crucial foundation for later development.^(1)^ However, many children grow up in environments that limit their developmental potential. Early intervention can significantly mitigate risks, but this depends on having access to appropriate tools for monitoring early child development (ECD) to identify children needing support at an early stage when brains are maximally plastic and responsive to change.

Many ECD assessments exist but their effectiveness and scalability are limited in resource-limited settings where, often, the need for such tools is greatest.^(2–4)^ Many tools (e.g., GMDS)^(5)^ are proprietary, require licensing fees, and must be administered by skilled professionals after extensive training. These requirements create geographic and socioeconomic inequities in access to appropriate screening and diagnostic tools, particularly in contexts where there is a paucity of professionals, most of whom are concentrated in more affluent urban areas. Moreover, because many aspects of development are influenced by culture, these tools are less applicable outside the Western contexts for which they were developed, and newer tools for non-Western settings often still depend on scarce specialists.^(6)^ Recent alternatives (e.g., Global Scales for Early Development; GSED^(7)^), address some of the limitations noted above, however, they are validated only up to age 3.

The STREAM platform was designed to overcome limitations of currently available measures through a mobile assessment administered by non-specialist workers (NSWs) with minimal training in low-resource community settings. Unlike tools relying solely on parent-report or observation, STREAM integrates parent-report, observation, and child performance on interactive tablet-based tasks, supporting scalability across diverse contexts. This study presents the first validation of STREAM in a large sample of children aged 0-6 years from New Delhi (India) and Blantyre (Malawi). We also generate reference curves to characterise age-related patterns of performance. These curves provide a framework for interpreting STREAM scores relative to age within this dataset (e.g., WHO growth standards^(8)^), but they are not intended as universal norms.

We aimed to harmonise the developmental assessments included in STREAM to generate domain-specific (motor/social/cognitive) scores and create age-standardised reference curves for these domains. We evaluated the structural, criterion, convergent, and known-groups validity of the scores, assessed their test-retest (TR) reliability, and examined their responsiveness to developmental change. In doing so, we test whether a digital, non-specialist delivered assessment can provide robust, context-appropriate measurement of neurodevelopment in two low-resource settings.

## Methods

### Study Design

The study design and protocol have been described in full. Briefly, this was a cross-sectional validation of STREAM with assessment of test-retest reliability and a longitudinal component to assess responsiveness. Data were collected between March 2022 and August 2024 in two diverse, low-resource, urban settings: New Delhi, India and Blantyre, Malawi. Ethical approval was obtained from local Institutional Review Boards (India: GD_2019_59; Malawi: P.01/21/3251), and informed caregiver consent was obtained. Assessments were conducted only when children were comfortable and stopped if they showed distress. COSMIN^(9)^ guidelines are used as a reporting standard and provided in Supplementary Materials 1.

### Sample

Children aged 0-6 years (Supplementary Materials 2 contains age-stratified sampling strategy) were recruited into either a *community* or an *enriched* sample (see ^(10)^ for eligibility criteria). The community sample included children recruited from antenatal or immunisation clinics and/or early education centres. The enriched sample comprised children referred/identified from tertiary hospitals and specialist clinics with existing or suspected neurodevelopmental conditions (e.g., autism spectrum disorder [ASD], global developmental delay [GDD], intellectual disability [ID]). This group was included to assess known-groups validity by testing how well STREAM distinguishes children with an elevated likelihood of neurodevelopmental conditions from those without. In India, enriched children had existing diagnoses via the All India Institute of Medical Sciences (AIIMS). In Malawi, diagnoses were less common, thus clinical officers used a structured proforma to document neurodevelopmental profiles and assign likely diagnoses. Supplementary Materials 3 summarises the enriched sample diagnoses, which were based on medical records in India and the proforma in Malawi.

### Measures

#### Primary Measures Assessment (PMA)

All children completed a core set of measures: STREAM (Figure 1), caregiver questionnaires, and anthropometry.

**Figure 1.**
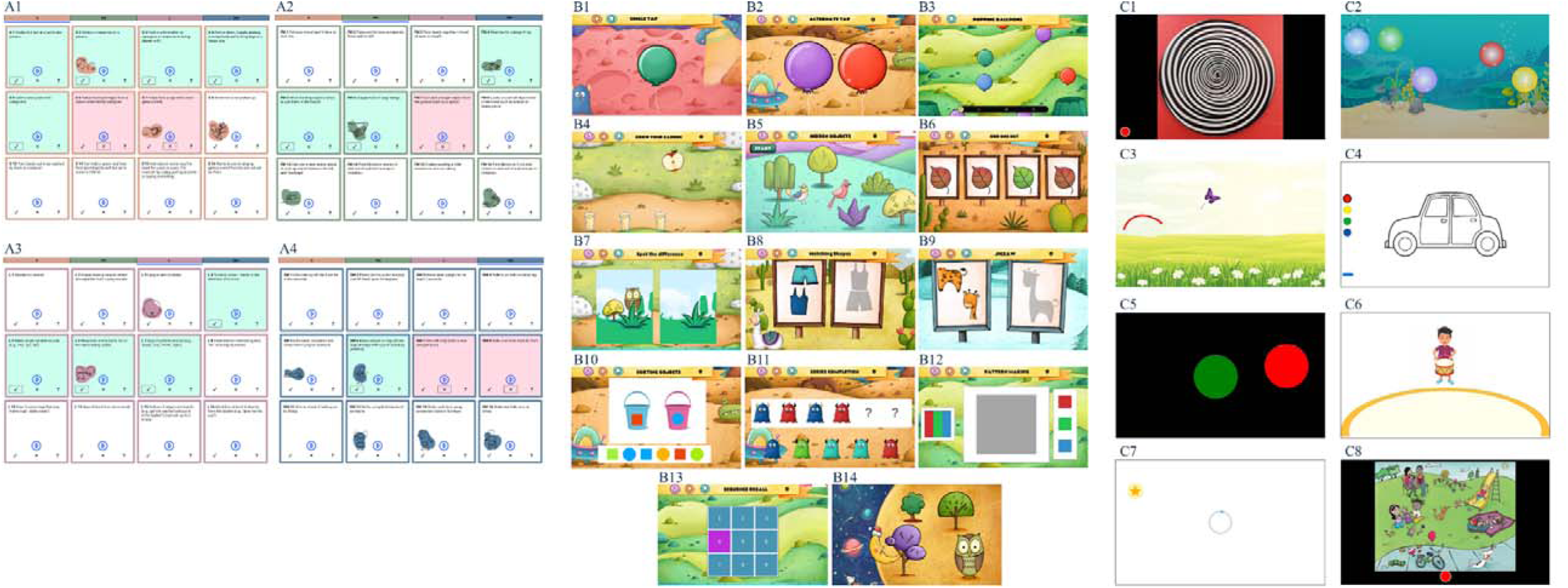
Visual depiction of each task within the STREAM platform. A: MDAT (a subset of items from each grid); A1: social; A2: fine motor; A3: language; A4: gross motor. B: DEEP; B1: single tap; B2: alternate tap; B3: popping balloons; B4: grow your garden; B5: hidden objects; B6: odd one out; B7: spot the difference; B8: matching shapes; B9: jigsaw; B10: sorting objects; B11: series completion; B12: pattern making; B13: sequence recall; B14: location recall. C: START; C1: wheel task; C2: bubble popping task; C3: motor following task; C4: colouring task; C5: button task; C6: synchrony task; C7: delayed gratification; C8: language sampling task. Figure 2 is based on Figure 2 in the published protocol.^(10)^ n.b. The images for the preferential looking task and parent-child interaction are not included in this figure.

##### STREAM

STREAM is a tablet-based application to assess ECD integrating content from three established tools: the Malawi Developmental Assessment Tool (MDAT^(11)^), DEvelopmental assessment on an E-Platform (DEEP^(12)^), and the Screening Tool for Autism Risk using Technology (START^(13)^). Culture-specific content from MDAT and START was reviewed with the original developers, and each tool was back-translated and piloted before integration into STREAM. Together, these tasks assess gross motor, fine motor, social, cognitive, and language abilities (Supplementary Materials 4). MDAT was administered to all children (0-6 years), and DEEP and most START tasks only to those aged 2.5-6 years who could interact independently with a tablet device. Two START components, the preferential looking task (PLT; social attention), and the Parent-Child Interaction (PCI; video-recorded play session), were administered to all ages. Metrics were derived according to each tool’s guidelines^(11–13)^ and are henceforth referred to as *STREAM-suite* metrics (Supplementary Materials 4).

##### Anthropometry

Height/length, weight, and head circumference were measured to derive WHO-standardised z-scores for weight-for-age (WAZ), height/length-for-age (HAZ), weight-for-length/height (WFL), and head circumference-for-age (HCZ).^(8)^

##### Parent/Caregiver questionnaires

Caregivers provided information on sociodemographic background and ECD risk-factors. Questionnaires included: Demographic and Health Survey (DHS^(14)^) for household background and socioeconomic status (SES); the Childhood Psychosocial Adversity Scale CPAS^(15)^) for exposure to early adversity; Family Care Indicators (FCI^(16)^) for home stimulation; Patient Health Questionnaire (PHQ-9^(17)^) for maternal depression; Child-Parent Relationship Scale (CPRS^(18)^)and Mother’s Object Relations Scale (MORS^(19)^) for caregiver-child relationships; Picture my Participation (PmP^(20)^) for the child’s participation in daily activities; and Brief Infant Sleep Questionnaire (BISQ^(21)^) for sleep quality. Caregivers also reported major life events (e.g., bereavement, displacement), child familiarity with digital technology, and developmental concerns using the Rashtriya Bal Swasthya Karyakram (RBSK^(22)^), a caregiver-report tool assessing multiple developmental domains (motor, language, cognitive, behaviour, attention).

#### Secondary Measures Assessment (SMA)

A subset of community children, selected with convenience sampling and stratified by age and sex, completed a *secondary assessment* within 3-4 working days of the PMA.

##### GMDS

The GMDS-3rd edition was administered to assess criterion validity of STREAM scores. Items were culturally and linguistically adapted at both sites in consultation with GMDS developers, and new reference data were derived jointly for India and Malawi as published UK norms were not representative (n.b. full details reported in a separate manuscript).

##### Hair cortisol

Hair samples (∼3cm) were collected, where consent and sample collection were possible, to measure hair cortisol concentration (HCC) as an exploratory biomarker of chronic stress over the past 2-3 months, given its established links with neurodevelopment.^(23)^ Samples from both sites were processed by Agilus Diagnostics Ltd (India) across 16 batches, each with a labelled and blinded duplicate. The mean coefficient of variation (7.2%) indicated acceptable assay consistency.

##### Braintools

As part of the wider SMA protocol, children completed the Braintools battery,^(24)^ combining EEG to measure brain responses to social/sensory stimuli with eye-tracking to capture visual attention. These data are beyond the scope of this paper and will be reported separately.

#### Test-retest

A convenience subsample stratified by age and sex was readministered STREAM after 7–10 days to evaluate reliability.

#### Longitudinal

A convenience subsample of children from the SMA, stratified by age and sex, were readministered STREAM, GMDS, and anthropometry after ∼18 months to assess responsiveness.

## Statistical Methods

Each STREAM tool (MDAT, DEEP, START) produces multiple metrics with distinct age-related trajectories (Figure 3). We applied Generalised Additive Models for Location, Scale, and Shape (GAMLSS) to derive age-standardised z-scores for these metrics for comparability. We combined these z-scores into STREAM domain scores (motor/social/cognitive) using Structural Equation Modelling (SEM^(25)^). Competing models were tested (Supplementary Figure 5) and fit evaluated with χ², RMSEA, CFI, TLI, and AIC. Metrics with loadings <0.34 were removed (Supplementary Table 4.2 includes list of included/excluded metrics). Cross-loadings (i.e., metrics loading onto more than one domain) suggested by modification indices were considered. A methods factor emerged which reflected systematic differences between tablet-based (DEEP, START) and non-tablet-based tasks (MDAT) and was retained in all models. STREAM domain scores were factor scores from the best-fitting SEM (Figure 4), age-standardised to provide reference curves for interpreting developmental performance relative to age *within our dataset*.

**Figure 3.**
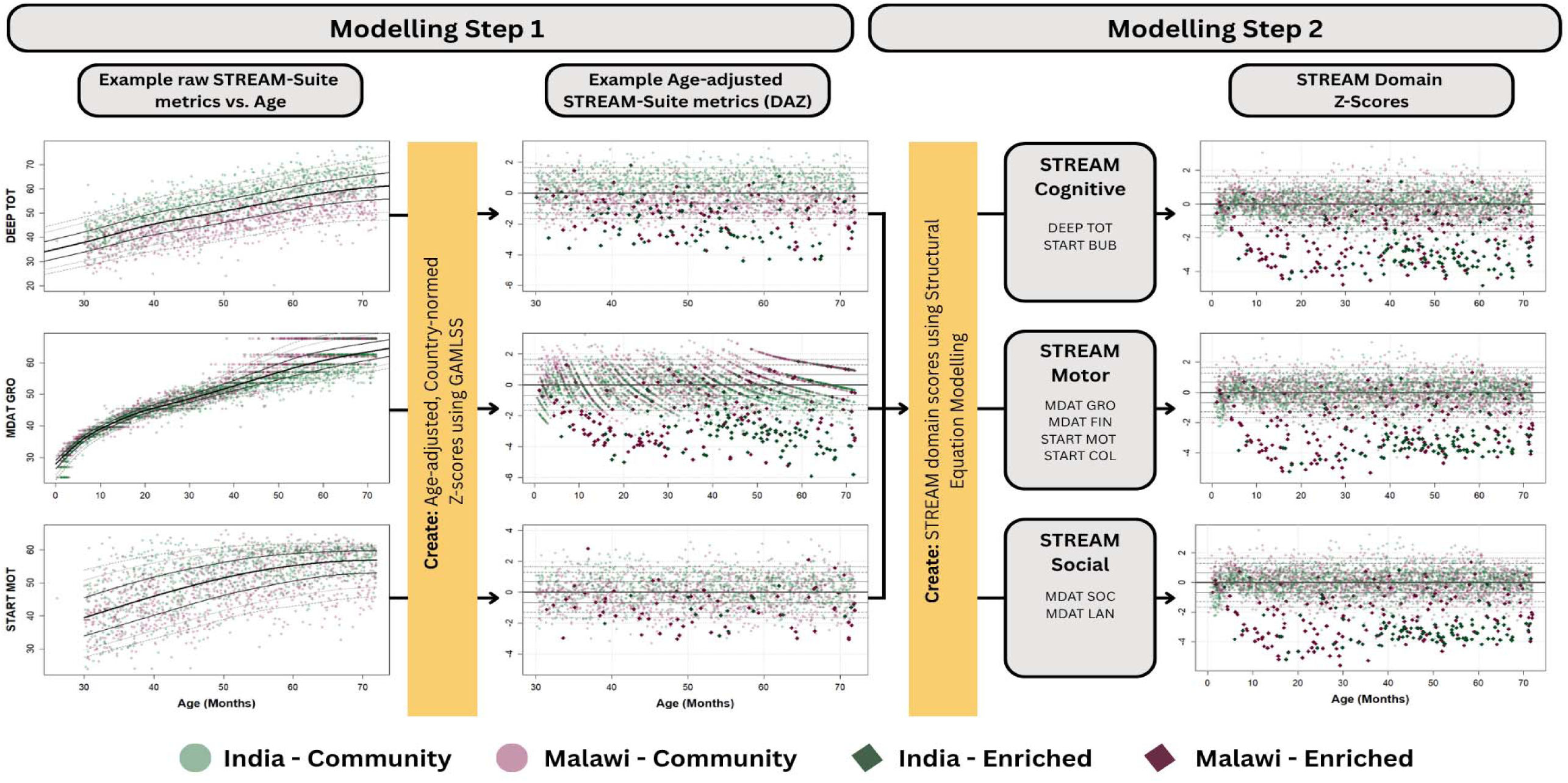
Overview of the modelling process with example metrics from each tool. Modelling step 1 applies GAMLSS to convert raw STREAM-suite metrics into age-adjusted z-scores. Modelling step 2 uses these z-scores as input to SEMs to derive latent STREAM domain scores (i.e., cognitive, motor, social) based on the pattern of loadings. These domain scores serve as country-specific reference curves. *MDAT FIN = MDAT Fine Motor; MDAT GRO = MDAT Gross Motor; START MOT = START Motor Following Task; START COL = START Colouring Task; START BUB = START Bubble Task; MDAT SOC = MDAT Social; MDAT LAN = MDAT Language*

**Figure 4.**
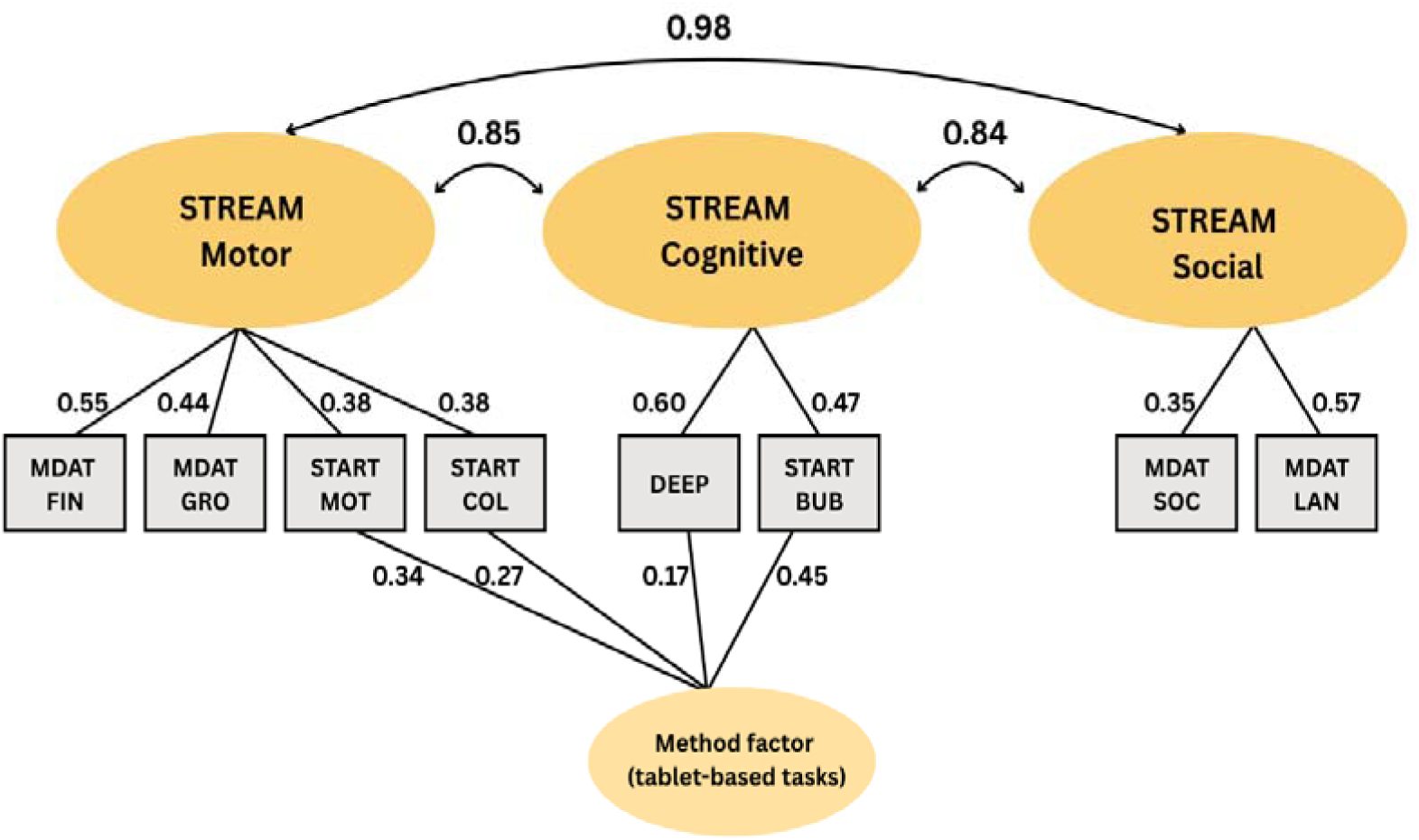
Model Path Diagram

Concurrent and convergent validity were assessed with Pearson’s or Spearman’s correlations (as appropriate). Known-groups validity was tested with AUC, assessing whether STREAM scores discriminate across: i) enriched vs community samples; ii) RBSK risk (<75% items passed [at-risk] vs >= 75% [not at-risk]); and iii) anthropometric risk (present vs absent). Test-retest reliability was evaluated using ICC (3,1) and responsiveness by correlating STREAM domain change scores with anthropometric measures and GMDS domains. All analyses were conducted in R 4.4.2.

### Missing Data

START and DEEP were administered only to children >2.5 years, creating *by-design* missingness. Our SEM addresses this via full-information maximum likelihood. As data were obtained through non-representative convenience and quota sampling, and additional participants were recruited to meet the quota when individuals declined to take part in the study, any efforts to reduce bias through missing data imputation would have limited utility.

### Sample size

Our planned cohort (N=4000) exceeded the SEM requirement of ∼1760 based on a 20:1 parameter-to-sample ratio.^(25)^ With n=4000, correlations of 0.10 and 0.30 yield one-sided 95% CI lower bounds of 0.07 and 0.27. For AUC (10% case prevalence; true AUC=0.80), the lower CI bound is 0.73. For test-retest (n=300; true reliability=0.80), the lower bound is 0.76.

## Results

Of 6580 families approached (community: 5866; enriched: 714), 6374 (97%) expressed interest (community: 5792; enriched: 582), and 6258 (98%) met eligibility criteria (community: 5753; enriched: 505; Figure 2). Among eligible families, 416 (7%) declined consent (community: 290; enriched: 126). Once enrolled targets were met, further eligible families were not enrolled.

**Figure 2.**
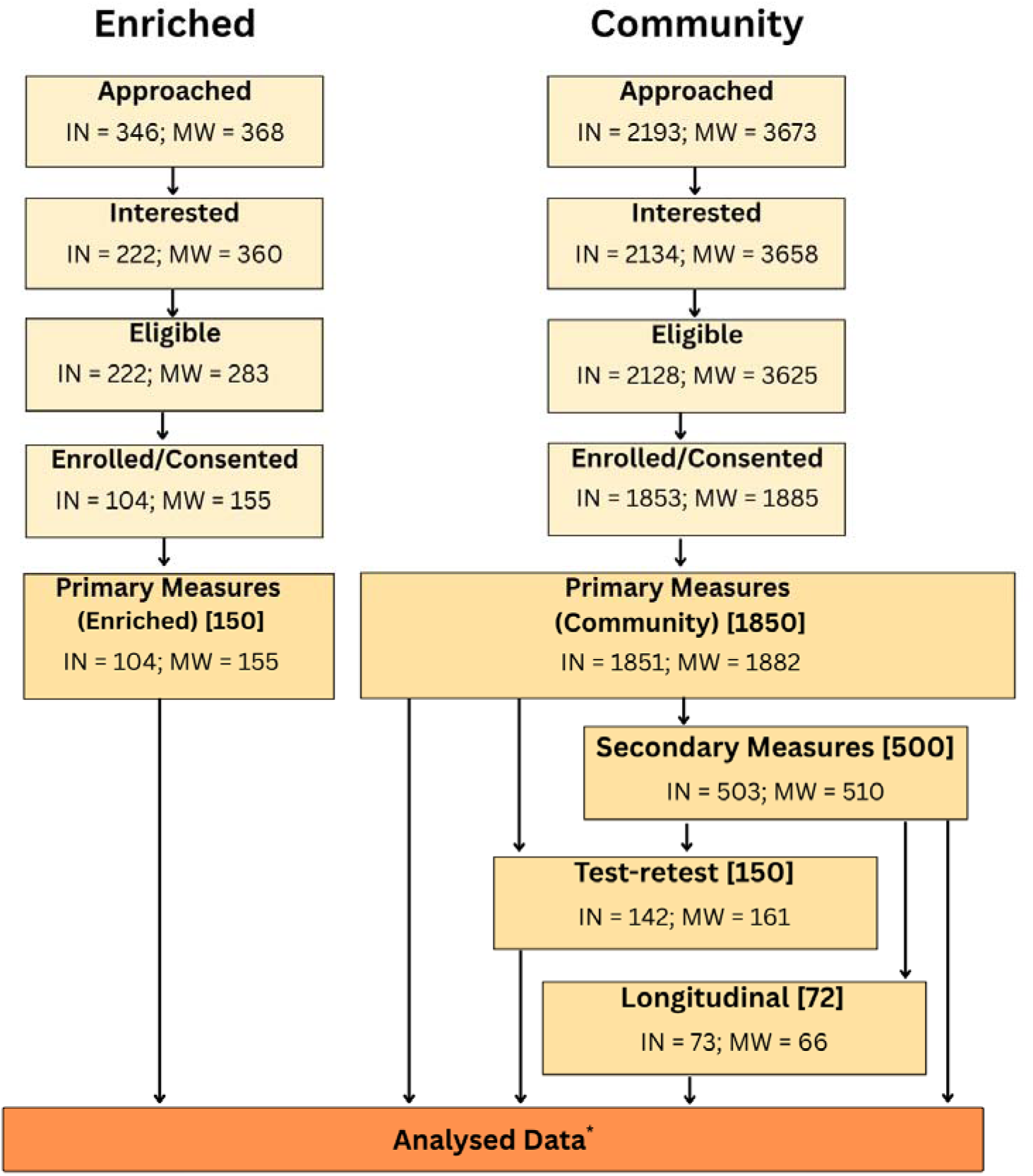
Flow diagram showing the number of children approached, eligible, enrolled, and assessed across key study components in India (IN) and Malawi (MW). *\*Specific sample sizes analysed in each component are detailed in Table 2. Target sample sizes for study components are reported in square brackets*.

In total, 3997 children were enrolled (community: 3738; enriched: 259). The PMA was completed by 3733 community and 259 enriched children. Test-retest data were available for 303 community children, and the SMA was administered to 1013 (all community), of whom 156 provided hair samples for HCC. The reduced hair sample size reflected budget constraints, parental refusal, and insufficient hair length. Of those in the SMA, 139 completed the longitudinal follow-up, lower than the planned 334 due to post-pandemic budget cuts.

Missing data was 2% for PMA, 1% for SMA, and 3% for longitudinal follow-up. A detailed analysis of the feasibility and acceptability of the STREAM tool is beyond the scope of the current paper and will be published separately.

### Sample Characteristics

Mean ages in the community samples were nearly identical across sites (India: 35.96 months, SD = 20.71; Malawi: 35.73 months, SD = 2.78; Table 1). Enriched children were older in India (46.75 months, SD = 15.56) than Malawi (36.55 months, SD = 20.03). The proportion of males was higher in enriched (India: 65.38%; Malawi: 61.94%) than community samples (India: 50.08%; Malawi: 49.84%).

**Table 1.**
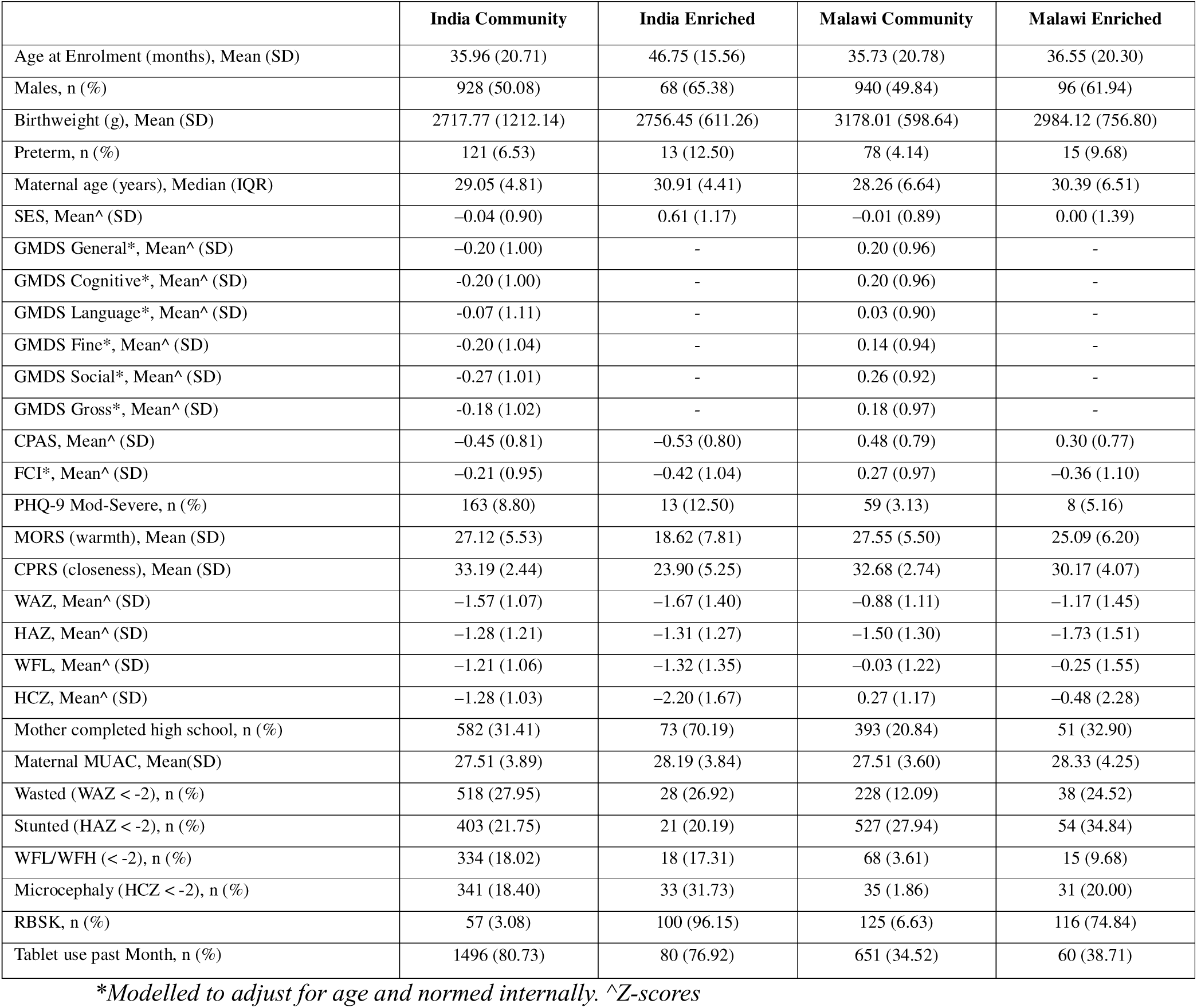
Sample Demographics and Scores.

**Table 2.** Psychometric properties of the STREAM Scores.

| Evidence type | Construct | Measure | Statistic | Sample size | India | Malawi | India | Malawi | India | Malawi | India | Malawi |
| --- | --- | --- | --- | --- | --- | --- | --- | --- | --- | --- | --- | --- |
|  |  |  |  |  | Achieved N |  | Motor |  | Attention & Cognition |  | Social & Communication |  |
| Criterion validity<br>(concurrent) | Child development | GMDS Total | Pearson | 1000 | 502 | 488 | **0.65**<br>(0.59-0.69) | **0.58**<br>(0.52-0.63) | **0.64**<br>(0.59-0.69) | **0.56**<br>(0.50-0.62) | **0.64**<br>(0.59-0.69) | **0.58**<br>(0.51-0.63) |
|  |  | GMDS Cognitive | Pearson | 1000 | 502 | 504 | **0.47**<br>(0.40-0.53) | **0.50**<br>(0.43-0.56) | **0.48**<br>(0.41-0.54) | **0.50**<br>(0.43-0.56) | **0.47**<br>(0.39-0.53) | **0.49**<br>(0.42-0.56) |
|  |  | GMDS Language | Pearson | 1000 | 502 | 494 | **0.55**<br>(0.48-0.61) | **0.42**<br>(0.34-0.49) | **0.56**<br>(0.49-0.61) | **0.40**<br>(0.33-0.48) | **0.55**<br>(0.49-0.61) | **0.42**<br>(0.35-0.49) |
|  |  | GMDS Fine | Pearson | 1000 | 502 | 498 | **0.50**<br>(0.44-0.57) | **0.51**<br>(0.44-0.57) | **0.50**<br>(0.43-0.56) | **0.49**<br>(0.42-0.56) | **0.50**<br>(0.43-0.56) | **0.50**<br>(0.44-0.57) |
|  |  | GMDS Social | Pearson | 1000 | 502 | 496 | **0.39**<br>(0.32-0.46) | **0.33**<br>(0.25-0.41) | **0.38**<br>(0.30-0.45) | **0.31**<br>(0.23-0.39) | **0.39**<br>(0.32-0.46) | **0.33**<br>(0.25-0.41) |
|  |  | GMDS Gross | Pearson | 1000 | 502 | 494 | **0.53**<br>(0.46-0.59) | **0.46**<br>(0.39-0.53) | **0.52**<br>(0.46-0.59) | **0.45**<br>(0.37-0.51) | **0.52**<br>(0.46-0.58) | **0.46**<br>(0.39-0.53) |
| Convergent validity | Household SES | DHS Wealth Index | Pearson | 4000 | 1853 | 1880 | **0.06**<br>(0.01-0.10) | **0.16**<br>(0.12-0.21) | **0.07**<br>(0.03-0.12) | **0.20**<br>(0.15-0.24) | **0.06**<br>(0.01-0.10) | **0.16**<br>(0.12-0.20) |
|  | Caregiver education | Mother's highest level of school attended | Spearman | 4000 | 1852 | 1880 | **0.12**<br>(0.08-0.17) | **0.15**<br>(0.11-0.2) | **0.13**<br>(0.09-0.18) | **0.18**<br>(0.13-0.22) | **0.12**<br>(0.08-0.17) | **0.15**<br>(0.11-0.19) |
|  |  | Father's highest level of school attended | Spearman | 4000 | 1842 | 1799 | **0.11**<br>(0.07-0.16) | **0.08**<br>(0.03-0.12) | **0.13**<br>(0.08-0.17) | **0.10**<br>(0.05-0.15) | **0.12**<br>(0.07-0.16) | **0.08**<br>(0.03-0.12) |
|  | Participation | PmP | Pearson | 2000 | 1078 | 1081 | **-0.14**<br>(-0.20--0.08) | **-0.15**<br>(-0.21--0.09) | **-0.12**<br>(-0.18--0.06) | **-0.11**<br>(-0.17--0.06) | **-0.14**<br>(-0.20--0.08) | **-0.15**<br>(-0.21--0.09) |
|  | Home stimulation | FCI | Pearson | 4000 | 1853 | 1884 | **0.25**<br>(0.20-0.29) | **0.20**<br>(0.15-0.24) | **0.24**<br>(0.20-0.29) | **0.18**<br>(0.14-0.22) | **0.25**<br>(0.20-0.29) | **0.20**<br>(0.15-0.24) |
|  | Parent-child relationship | MORS (0-3 years) (warmth) | Pearson | 2000 | 930 | 916 | **0.19**<br>(0.12-0.25) | **0.11**<br>(0.05-0.17) | **0.19**<br>(0.12-0.25) | **0.11**<br>(0.05-0.18) | **0.19**<br>(0.12-0.25) | **0.11**<br>(0.04-0.17) |
|  |  | CPRS (3-6 years) (closeness) | Pearson | 2000 | 924 | 852 | **0.34**<br>(0.29-0.40) | 0.06<br>(0.00-0.13) | **0.34**<br>(0.28-0.40) | 0.09<br>(0.02-0.15) | **0.35**<br>(0.29-0.40) | 0.06<br>(-0.01-0.13) |
|  |  | PCI+ (scaled score) | Pearson | 4000 | 287 | 270 | **0.26**<br>(0.15-0.37) | 0.04<br>(-0.08-0.16) | **0.26**<br>(0.15-0.37) | 0.03<br>(-0.09-0.15) | **0.26**<br>(0.15-0.37) | 0.04<br>(-0.08-0.16) |
|  | Exposure to conflict; abuse | CPAS | Pearson | 4000 | 1853 | 1765 | -0.02<br>(-0.06-0.03) | **-0.10**<br>(-0.15--0.05) | -0.03<br>(-0.07-0.02) | **-0.10**<br>(-0.15--0.05) | -0.02<br>(-0.06-0.03) | **-0.10**<br>(-0.15--0.06) |
|  | Caregiver depression | PHQ-9 | Pearson | 4000 | 1853 | 1765 | -0.02<br>(-0.06-0.03) | -0.03<br>(-0.08-0.01) | -0.02<br>(-0.07-0.02) | -0.03<br>(-0.07-0.02) | -0.02<br>(-0.06-0.03) | -0.03<br>(-0.08-0.01) |
|  | Sleep quality | BISQ | Pearson | 4000 | 346 | 865 | -0.07<br>(-0.17-0.04) | -0.05<br>(-0.11-0.02) | -0.07<br>(-0.18-0.03) | -0.05<br>(-0.12-0.02) | -0.07<br>(-0.17-0.04) | -0.05<br>(-0.11-0.02) |
|  | Exposure to stress | Hair cortisol | Pearson | 200 | 106 | 50 | -0.12<br>(-0.30-0.08) | 0.18<br>(-0.11-0.43) | -0.11<br>(-0.30-0.08) | 0.21<br>(-0.07-0.46) | -0.12<br>(-0.30-0.08) | 0.17<br>(-0.11-0.43) |
| Known Groups<br>(Convergent) | Neurodevelopmental risk | RBSK Cut score | AUC | 4000 | 1922 | 1991 | **0.93**<br>**(0.91-0.96)** | **0.84**<br>**(0.81-0.87)** | **0.93**<br>**(0.91-0.95)** | **0.83**<br>**(0.80-0.87)** | **0.93**<br>**(0.91-0.96)** | **0.84**<br>**(0.81-0.87)** |
|  | Wasting | WAZ<-2 | AUC | 4000 | 1584 | 1663 | **0.79**<br>**(0.71-0.86)** | **0.71**<br>**(0.62-0.80)** | **0.78**<br>**(0.71-0.86)** | **0.71**<br>**(0.62-0.79)** | **0.79**<br>**(0.71-0.86)** | **0.71**<br>**(0.62-0.80)** |
|  | Stunting | HAZ<-2 | AUC | 4000 | 1597 | 1698 | **0.78**<br>**(0.70-0.85)** | **0.70**<br>**(0.62-0.78)** | **0.77**<br>**(0.69-0.84)** | **0.70**<br>**(0.62-0.78)** | **0.78**<br>**(0.71-0.86)** | **0.70**<br>**(0.62-0.78)** |
|  | Malnutrition | WHZ<-2 | AUC | 4000 | 1578 | 1658 | **0.68**<br>**(0.58-0.79)** | **0.59**<br>**(0.48-0.71)** | **0.68**<br>**(0.57-0.79)** | **0.60**<br>**(0.49-0.71)** | **0.68**<br>**(0.57-0.79)** | **0.60**<br>**(0.48-0.71)** |
|  | Microcephaly | HCZ<-2 | AUC | 4000 | 1597 | 1699 | **0.82**<br>**(0.74-0.90)** | **0.75**<br>**(0.64-0.85)** | **0.82**<br>**(0.74-0.89)** | **0.74**<br>**(0.64-0.85)** | **0.82**<br>**(0.74-0.90)** | **0.75**<br>**(0.64-0.85)** |
|  | Sample | Enriched Vs Community | AUC | 4000 | 1957 | 2041 | **0.98**<br>**(0.96-1.00)** | **0.90**<br>**(0.87-0.94)** | **0.97**<br>**(0.95-0.99)** | **0.89**<br>**(0.86-0.93)** | **0.98**<br>**(0.96-1.00)** | **0.90**<br>**(0.87-0.94)** |
| Responsiveness | Child development | GMDS Total | Pearson | 144 | 69 | 63 | **0.26**<br>**(0.01-0.47)** | 0.10<br>(-0.16-0.34) | 0.05<br>(-0.19-0.29) | 0.14<br>(-0.11-0.38) | 0.20<br>(-0.04-0.43) | 0.10<br>(-0.16-0.34) |
|  |  | GMDS Cognitive | Pearson | 144 | 69 | 65 | 0.16<br>(-0.08-0.39) | 0.06<br>(-0.19-0.31) | 0.05<br>(-0.19-0.28) | 0.12<br>(-0.13-0.35) | 0.12<br>(-0.13-0.35) | 0.06<br>(-0.19-0.31) |
|  |  | GMDS Language | Pearson | 144 | 69 | 63 | **0.35**<br>**(0.11-0.54)** | 0.09<br>(-0.16-0.34) | 0.19<br>(-0.05-0.41) | 0.09<br>(-0.17-0.33) | **0.32**<br>**(0.09-0.53)** | 0.08<br>(-0.17-0.33) |
|  |  | GMDS Fine | Pearson | 144 | 69 | 64 | 0.01<br>(-0.23-0.26) | 0.07<br>(-0.18-0.31) | -0.16<br>(-0.38-0.08) | 0.06<br>(-0.19-0.30) | -0.03<br>(-0.27-0.22) | 0.07<br>(-0.18-0.32) |
|  |  | GMDS Social | Pearson | 144 | 69 | 63 | 0.21<br>(-0.04-0.43) | 0.01<br>(-0.24-0.26) | 0.10<br>(-0.14-0.33) | 0.08<br>(-0.17-0.32) | 0.20<br>(-0.05-0.42) | 0.01<br>(-0.24-0.26) |
|  |  | GMDS Gross | Pearson | 144 | 69 | 64 | 0.12<br>(-0.13-0.36) | 0.09<br>(-0.16-0.34) | 0.04<br>(-0.2-0.28) | 0.14<br>(-0.11-0.38) | 0.11<br>(-0.14-0.34) | 0.09<br>(-0.16-0.34) |
|  | Wasting | WAZ | Pearson | 144 | 53 | 52 | 0.21<br>(-0.07-0.46) | 0.00<br>(-0.27-0.28) | 0.13<br>(-0.14-0.39) | 0.07<br>(-0.2-0.34) | 0.2<br>(-0.08-0.45) | 0.00<br>(-0.28-0.27) |
|  | Stunting | HAZ | Pearson | 144 | 53 | 52 | 0.22 | 0.24 | 0.16 | 0.28 | 0.22 | 0.24 |
|  |  |  |  |  |  |  | (-0.06-0.47) | (-0.04-0.48) | (-0.11-0.42) | (0.00-0.51) | (-0.06-0.46) | (-0.04-0.48) |
|  | Malnutrition | WHZ | Pearson | 144 | 53 | 53 | 0.10<br>(-0.18-0.37) | -0.24<br>(-0.48-0.04) | 0.05<br>(-0.23-0.32) | -0.22<br>(-0.46-0.06) | 0.10<br>(-0.18-0.37) | -0.25<br>(-0.49-0.03) |
|  | Microcephaly | HCZ | Pearson | 144 | 53 | 52 | 0.21<br>(-0.07-0.46) | -0.02<br>(-0.29-0.26) | 0.17<br>(-0.11-0.42) | 0.06<br>(-0.22-0.33) | 0.2<br>(-0.08-0.45) | -0.02<br>(-0.30-0.26) |
| **Test-retest reliability** |  | STREAM Scores | ICC (3,1) | 300 | 138 | 148 | **0.69**<br>**(0.59-0.73)** | **0.67**<br>**(0.57-0.75)** | **0.65**<br>**(0.55-0.74)** | **0.45**<br>**(0.31-0.56)** | **0.69**<br>**(0.59-0.77)** | **0.66**<br>**(0.56-0.75)** |

<sup>^</sup>Anthropometric Z-scores were calculated only for children aged 0–5 years. This is because WHO growth standards differ for children under and over 5 years: the 0–5-year standards are based on a multi-country, optimal sample and are widely validated, whereas the 5+ standards are based on data from a single country and are less robust. As a result, Z-scores cannot be consistently applied across the full age range, and we restricted analyses to children under 5 to ensure comparability and validity. +We derived a summary score for PCI using a graded response factor model. Codes for attention to caregiver, effective communication, and infant–caregiver engagement showed strong factor loadings (0.95, 0.89, and 0.91, respectively). Positive and negative affect loaded modestly (0.58 and –0.43) and liveliness had a low loading (0.27). Although all children participated in the PCI task, the analytic sample is currently small due to feasibility constraints, as video coding is conducted manually by trained raters.

Community samples also differed between sites. For all reported z-scores, higher values indicate better outcomes. Malawian children had higher GMDS z-scores (range [0.03,0.26]) than Indian children (range [-0.27,0.07]), as well as higher CPAS (0.48 vs -0.45) and FCI z-scores (0.27 vs -0.21). Mothers in Malawi had lower rates of moderate-to-severe maternal depression (PHQ-9; 3.13% vs 8.80%) and lower child tablet use in the past month (38.71% vs 80.73%).

Across both countries, enriched children differed from community children; they were more often born preterm (India: 12.50% vs 6.53%; Malawi: 9.68% vs 4.14%), had lower anthropometric indices, and more frequently flagged as at-risk on the RBSK (India: 96.15% vs 3.08%; Malawi: enriched = 74.84%, community = 6.63%). In India, the enriched sample also showed unexpected differences, with higher SES z-scores (0.61 vs -0.04) and a greater proportion of mothers completing high school (70.19% vs 31.41%), likely reflecting recruitment through a tertiary hospital. These differences were not observed in Malawi.

### Model Fit and Structural Validity

Several competing models were tested (Supplementary Materials 5); no models with cross-loadings were theoretically justified based on modification indices. The final, best-fitting model (Model 2) used to generate STREAM domain scores is shown in Figure 4. Model 2 (Table 2) exceeded conventional fit benchmarks (RMSEA=0.03; CFI=0.98; TLI=0.97). Although the chi-square test was significant (χ²LL=58.63, p<0.001), this is common in large samples and does not indicate poor fit.^(25)^ Mean standard errors of measurement were 0.55 (SD = 0.04) for motor, 0.64 (SD = 0.07) for cognitive, and 0.56 (SD = 0.04) for social domains. Compared with a unidimensional model including only a ‘total’ score rather than separate domain scores, Model 2 showed a negligible AIC improvement, suggesting STREAM scores lie near the boundary between uni- and multi-dimensionality. Excluding the methods factor led to convergence issues due to high inter-domain correlations. Even with the factor included, correlations remained high, indicating that domains can be estimated separately but are substantially overlapping. Final STREAM domain scores, derived from Model 2, provide the age-standardised reference curves.

### Psychometric Properties

Criterion validity was supported by correlations between STREAM and GMDS domains (India: range [0.38,0.65]; Malawi: range [0.31,0.58]; Table 2).

Convergent validity varied by measure. STREAM scores correlated with FCI (range [0.18,0.29]) and the warmth subscale of MORS (range [0.11,0.25]). Smaller correlations were seen with maternal education (range [0.12,0.18]), PmP (range [-0.15,-0.11]), DHS Wealth Index (SES; range [0.06,0.20]), and paternal education (range [0.08,0.13]). Country-specific findings included correlations with CPRS (range [0.34,0.35]) and PCI in India (all STREAM domains *r* = 0.26) and CPAS in Malawi (all STREAM domains *r* = -0.10). No corelations were found with maternal depression (PHQ-9), sleep quality (BISQ), or HCC.

Known-groups validity was strong. STREAM scores discriminated i) enriched vs community children (India: AUC range [0.90,0.98]; Malawi: AUC range [0.89,0.90]), ii) children flagged as at-risk vs not at-risk on RBSK (India: AUC = 0.93; Malawi: AUC = 0.84), and iii) children with and without anthropometric risk: those with stunting/wasting (AUC > 0.70) and those with head circumference 2SDs below the mean (India: AUC = 0.82; Malawi: AUC = 0.74). These analyses support that STREAM distinguishes groups where developmental differences would be expected.

Test-retest reliability was moderate (India: range [0.65,0.69]; Malawi: range [0.45,0.67]), with lowest reliability in the Malawi cognitive domain (0.45).

Responsiveness analyses indicated no longitudinal correlations in Malawi. In India, change in STREAM motor scores correlated with GMDS total (0.26) and language (0.35), and change in STREAM social scores with GMDS language (0.32). No correlations were found between changes in STREAM scores and anthropometry.

## Discussion

The STREAM platform is a non-specialist, tablet-based approach for scalable assessment of ECD in children aged 0-6 years. It addresses key recommendations highlighted by recent reviews (e.g.,^(4)^); it spans the full early childhood period, covers motor, social, and cognitive domains, and combines direct child assessment, caregiver-report, and observational measures for use in low-resource community settings. We evaluated STREAM’s psychometric properties, including structural, criterion, convergent, and known-groups validity, as well as test-retest reliability and responsiveness to change. Findings provide evidence for STREAM’s validity as a measure of ECD in India and Malawi, and highlight its potential to reduce inequities in access to child neurocognitive assessment tools where specialist tools are scarce. The domain-specific reference curves generated also provide a resource for benchmarking and monitoring child development in these settings.

Criterion validity was robust, with strong correlations between age-adjusted STREAM and GMDS scores (*r* = 0.56-0.65). Known-groups analyses, which were conducted to test whether STREAM scores distinguished between groups where developmental differences would be expected, was also supported. STREAM distinguished i) enriched from community samples, ii) children at-risk vs not at-risk on the RBSK, and iii) those with anthropometric risk such as stunting, wasting, or head circumference 2SDs below the mean. These results align with well-established evidence linking early growth faltering to poorer developmental outcomes^(26)^ and reinforce STREAM’s validity in distinguishing groups with developmental risk, supporting its use for early identification and referral.

Convergent validity was supported by correlations with relevant developmental constructs, most in the expected direction though not all statistically significant. The strongest correlations were with the CPRS Closeness subscale (ages 3-6 years) in India (*r* = 0.34-0.35), consistent with prior links between parent-child closeness/sensitivity and early cognitive and socio-emotional outcomes.^(27)^ This pattern was not seen in Malawi, possibly reflecting cultural differences in how closeness is experienced, expressed, or reported, or in parental responses to developmental delays. In contrast, the MORS warmth subscale (0-3 years), which assesses similar constructs in younger children, showed significant correlations at both sites. This may suggest that cultural differences in expressing closeness or affection become more pronounced or context-dependent as children grow older, affecting both the construct and the sensitivity of the CPRS in Malawi.

PCI scores correlated with all STREAM domains in India (r = 0.26) but not Malawi. The PCI score is a composite measure driven by factors relevant to the parent-child relationship, such as child attention to caregiver, effective child communication, and infant-caregiver engagement. Its lack of correlation in Malawi may suggest that the task does not capture culturally typical parent-child interaction.^(28)^ Indeed, many Malawian caregivers reported that the PCI’s ‘free play’, which involves one-to-one play with toys in front of a camera followed by a period of instructed disengagement from the child, felt unnatural and unrepresentative of home practices. Taken together, these findings highlight the influence of cultural norms on the expression and reporting of parent-child interactions and the importance of adapting and validating measures across contexts.

Correlations with other convergent measures were smaller but generally consistent with theoretical expectations. Across sites, higher STREAM scores were associated with greater home learning stimulation (FCI: range [0.18,0.25]) and higher maternal education (range [0.12, 0.18]), consistent with evidence on the role of the home environment in early development.^(1)^ Weaker associations were observed with paternal education and SES. A negative correlation with PmP scores suggests that children with higher STREAM scores may encounter fewer barriers to daily activities.^(29)^ Although effect sizes were small, the direction and consistency of correlations with convergent measures were comparable to prior studies,^(30)^ supporting the validity of STREAM scores in capturing meaningful variation in children’s development.

Systematic site differences were observed. Malawian children scored lower overall than Indian children on tablet-based tasks (DEEP and some START tasks), possibly reflecting lower digital familiarity, as recent tablet use was substantially higher in India (76.9% – 80.7%) than in Malawi (34.5% – 38.7%). All children completed a short touchscreen familiarisation task to practise basic actions such as tapping, dragging, and dropping, but this may not have fully mitigated differences in prior exposure. In contrast, Malawian children scored higher on the GMDS across domains, and site differences were smaller for observational and caregiver-report measures (e.g., MDAT). As the GMDS is not designed for cross-country comparability, our analyses focus on within-country correlations rather than pooled estimates with overall bias less relevant to our validation evidence.

Test-retest reliability indicated that STREAM scores were generally stable. Although ICC (2,1) (absolute agreement) was originally planned, ICC (3,1) (consistency) was used to account for systematic differences between sessions arising from *learning effects* on tablet-based tasks. These effects were likely amplified in Malawi, where digital familiarity was lower. Reliability for the attention and cognition domain, driven entirely by tablet-based tasks, was lower in Malawi (0.45) compared with other domains across both sites (range [0.65, 0.69]). This lower reliability likely reflects variable baseline performance in children with less tablet experience, leaving more scope for improvement between sessions and reducing correlation across timepoints.

Responsiveness analyses were constrained by a small longitudinal sample (n = 139 vs planned 334), due to post-pandemic funding cuts, reducing power from 0.80 to 0.40. Significant correlations were observed in India between change in STREAM motor scores and GMDS total (0.26), and between GMDS language scores and STREAM motor (0.35) and social (0.32) domains. Other correlations were non-significant, including with anthropometry. These limited findings likely reflect low power rather than insensitivity of the STREAM tool. Further research is needed to assess its ability to capture developmental change over time.

Limited differentiation was observed between motor, cognitive, and social domain scores, particularly in younger children. Both score patterns (Figure 3) and SEM analyses (Figure 4) showed substantial inter-domain overlap. Although domain-specific models fit adequately, a single-factor model performed comparably, and this convergence persisted across age groups. These findings align with prior reports of domain specific scores being of questionable utility in younger children(31) and provide critical insights about the added value of domain-specific vs general developmental constructs. This overlap may partly reflect the inherent challenge of designing tasks for young children that isolate single domains, as most require shared abilities (e.g., attention, language, social interaction). While this overlap is expected, it does not preclude the possibility that children with specific developmental difficulties may show uneven profiles (e.g., strong cognition but weak social skills), which warrants further investigation.

Several tasks were excluded from final domain scores due to low factor loadings during SEM fitting (Supplementary Table 4.2). However, tasks such as the PLT (social preference/attention) and Wheel Task (sensory seeking) distinguished ASD from non-ASD children (results to be reported separately), suggesting they may provide valuable information even though they do not map cleanly onto broad domains. Future work could develop composite indicators to capture such functional areas (e.g., sensory or social functioning) for use in monitoring and referral.

Ongoing work uses machine-learning to optimise the STREAM battery by identifying the most informative tasks for different clinical use cases. This approach aims to reduce administration time while preserving psychometric performance. The app’s modular structure already allows assessors to select relevant components for specific contexts, reducing burden and improving feasibility in low-resource and longitudinal studies.

In summary, the findings support STREAM as a valid and reliable tool for assessing ECD in low-resource settings. The platform demonstrated robust psychometric properties, and country-specific reference curves improve the interpretability of scores in India and Malawi. While this paper focuses on psychometric validation, it is important to note that STREAM’s design incorporates features that support scalability and practical use in low-resource settings, including tablet-based administration, automated scoring of assessments, offline functionality, and suitability for use by non-specialist workers. Together, these features may facilitate broader uptake of the tool, both in research settings and in public health programmes, and help reduce current inequities in access to ECD assessment.

## Data Availability

All data produced in the present study are available upon reasonable request to the authors

## Data Sharing Statement

Deidentified participant data and a relevant data dictionary will be made available for this study on an institutional data repository on publication of the paper.

## Author contribution statement

All authors contributed substantially to this work. EHW, GM and NMT led the drafting of the manuscript. GM, EHW, AR, and NMT performed the data analysis with contributions from BC, SB, and SC. BC, MG, GD, EM, SC, VP, EJ, MKB, MHJ, GM, DM and SB led the conceptualisation of the study. MMCL, SB, DG, TDB, GLE, LM, VN, CN, RN, AB, AR, UK, NM, AS, IM, AC, SG, DM and EM contributed to on-site implementation and drafting of site-specific or tool-specific details for the manuscript. All authors reviewed and edited the manuscript and read and approved the final version prior to submission.

