## Supplementary Materials for "Comprehensive neurodevelopmental assessment through non-specialists: Validation of the STREAM digital platform in India and Malawi"

### Supplementary Materials 1 – Cosmin Reporting Checklist

The process of validation involves gathering appropriate evidence to support score interpretation. We use the COSMIN reporting guidelines for studies on measurement properties of patient-reported outcome measures (Gagnier, de Arruda, Terwee, & Mokkink, 2025).

Supplementary Table 1. Cosmin reporting checklist

| **Item number – item name** | **Item** | **Evidence** |
| --- | --- | --- |
| **Report section: Title** |  |  |
| T1 – Title | Identify the report as a study of one or more measurement properties of a specific PROM to measure a specified construct in a specified population. | Comprehensive neurodevelopmental assessment through non-specialists: Validation of the STREAM digital platform in India and Malawi |
| **Report section: Abstract** |  |  |
| A1 – Objectives | Provide the specific objective(s) of the research, specifying:  (1) the name (and version, if relevant), and construct(s) of the PROM,  (2) the measurement properties being evaluated, and  (3) relevant study characteristics. | (1) *“The Scalable TRansdiagnostic Early Assessment of Mental health (STREAM)”,*  (2) *“Motor, cognitive, and social domain scores were derived from age-adjusted data using structural equation modelling (SEM). Validity was assessed against the Griffiths Mental Development Scales (GMDS), caregiver-reported developmental measures, and anthropometric indices. We examined known-groups discriminability, test-retest reliability, and 18-month responsiveness to change.”*    (3) *“[STREAM] … uses a mobile platform, delivered by non-specialist workers, to assess motor, social, and cognitive development in children aged 0-6 years”* |
| A2 – Design | Specify (details of the) study design used to evaluate the measurement properties. | By their nature, convergent and concurrent validity are cross-sectional, and responsiveness is most closely related to longitudinal. Test-retest reliability is a study type of its own and it would be difficult to say it was *longitudinal*. That leaves structural validity, which in this case was cross-sectional, and Known groups, which was also cross-sectional. However, the importance of the study design for these validity types is not important enough to mention in the abstract. |
| A3 – Methods | Specify the methods for evaluating each measurement property. | Structural Validity – SEM  Criterion Validity – Pearson’s/Spearman’s  Convergent Validity – Pearson’s/Spearman’s  Known-groups Validity - AUC  Test-retest reliability - ICC  Responsiveness – Pearson’s/Spearman’s on change scores |
| A4 – Results | Provide the main results for all measurement properties evaluated. | See Findings Section in Abstract. |
| A5 – Discussion/Conclusions | Provide a brief statement of the implications of the findings in the context of existing evidence on the PROM. | This is the first study to evaluate the PROM, as such there is no previous evidence |
| **Report section: Introduction** |  |  |
| I1 – PROM | Specify the name and, if relevant, the version, and construct(s) of the PROM. | *“Comprehensive neurodevelopmental assessment through non-specialists: Validation of the STREAM digital platform in India and Malawi”s* |
| I2 – Target population | Specify the target population and context of use that the PROM was designed for. | *“This study presents the first validation of STREAM in a large sample of children aged 0-6 years from New Delhi (India) and Blantyre (Malawi).”* |
| I3 – State of knowledge & Rationale | Provide a description of the current scientific knowledge (what is known and not known) regarding the measurement properties of the PROM. Explain why the new study is necessary. Provide citations for the original development paper(s). | As the PROM was created in this paper this is not relevant. |
| I4 – Objectives | Provide the specific objective(s) of the research, specifying  (1) the name (and version, if relevant) of the PROM,  (2) the measurement properties being evaluated, and  (3) relevant study sample characteristics. | (1) *“The Scalable TRansdiagnostic Early Assessment of Mental health (STREAM)”,*  (2) *“Motor, cognitive, and social domain scores were derived from age-adjusted data using structural equation modelling (SEM). Validity was assessed against the Griffiths Mental Development Scales (GMDS), caregiver-reported developmental measures, and anthropometric indices. We examined known-groups discriminability, test-retest reliability, and 18-month responsiveness to change.”*    (3) *“[STREAM] … uses a mobile platform, delivered by non-specialist workers, to assess motor, social, and cognitive development in children aged 0-6 years”* |
| **Report section: General Methods** |  |  |
| GM1 – Study design | Specify (details of the) study design used to evaluate the measurement properties. | *“Briefly, this was a cross-sectional validation of STREAM with assessment of test retest-reliability and a longitudinal component to assess responsiveness.”* |
| GM2 – Participants | Specify how the study participants were selected. Specify the inclusion and exclusion criteria | See protocol paper:  Williams EH, Thompson NM, McCray G, Crespo-Llado MM, Bhavnani S, Gajria D, et al. Scalable Transdiagnostic Early Assessment of Mental Health (STREAM): a study protocol. BMJ Open. 2024 June 1;14(6):e088263  “Children will be eligible for STREAM if:  ►They are between 0 and 72 months of age (ie, 0–6 years).  ►Their parent/caregiver can provide informed consent.  ►They and their parent/caregiver reside within the catchment areas of the study sites.    Children will be excluded if:  ►Their sibling has participated in the STREAM study.  ►They have a severe vision, hearing or motor impairment, as  reported by their  parent/caregiver, which  would limit their ability to interact with a tablet device.  ►They have had an uncontrolled seizure in the last 48 hours that lasted more than 5 min.  ►They are currently enrolled in another research study or trial.  ►Their parent/caregiver has a severe vision or hearing impairment.  ►Their parent/caregiver has a severe learning disability or a current, severe psychiatric condition. |
| GM3 – PROM details | Provide details about the original version of the PROM as well as of the PROM version being studied, specify the conceptual framework (reflective/formative model), details on the structure (the number of items and subscales), the language, response scale, recall period, direction of scoring, and scoring algorithm of the PROM. Specify how the PROM was administered (e.g., in what setting, mode of administration (e.g. paper, electronic) what instructions were given), including the country in which it is administered | As this is a new PROM there is no original version but it is based on three existing tools. The construction and validation of these tools can be found in;  *Gladstone M, Lancaster GA, Umar E, Nyirenda M, Kayira E, Van Den Broek NR, et al. The Malawi Developmental Assessment Tool (MDAT): The Creation, Validation, and Reliability of a Tool to Assess Child Development in Rural African Settings. Osrin D, editor. PLoS Med. 2010 May 25;7(5):e1000273.*  *Bhavnani S, Ranjan A, Mukherjee D, Divan G, Prakash A, Yadav A, Lal C, Gajria D, Irfan H, Sharma KK, Todkar S, Patel V, McCray G. A non-specialist worker delivered digital assessment of cognitive development (DEEP) in young children: A longitudinal validation study in rural India. PLOS Digit Health. 2025 May 16;4(5):e0000824. doi: 10.1371/journal.pdig.0000824. PMID: 40378360; PMCID: PMC12084064.*  *Dubey I, Bishain R, Dasgupta J, Bhavnani S, Belmonte MK, Gliga T, et al. Using mobile health technology to assess childhood autism in low-resource community settings in India: An innovation to address the detection gap. Autism Int J Res Pract. 2024 Mar;28(3):755–69.*  The remaining information is detailed throughout the paper. |
| GM4 – Additional data collection | Describe why and how other data was collected (e.g., construct and measurement properties of the comparator instruments, characteristics of groups being compared, and rationale for choosing groups), including mode of administration (e.g., paper, electronic). | See Methods Section. |
| GM5 – Time points procedures | Provide all time points of all measurements. | See Methods Section. |
| GM6 – Justification for sample size | Provide a rationale for the sample size for all measurement properties analyses (including subgroups). | *“Our planned cohort (N=4000) exceeded the SEM requirement of ~1760 based on a 20:1 parameter-to-sample ratio.(25) With n=4000, correlations of 0.10 and 0.30 yield one-sided 95% CI lower bounds of 0·07 and 0·27. For AUC (10% case prevalence; true AUC=0·80), the lower CI bound is 0·73. For test-retest (n=300; true reliability=0·80), the lower bound is 0·76.”* |
| GM7 – Statistical analyses | Describe the statistical analyses corresponding to all objectives (see measurement properties specific boxes).  Describe the criteria for good measurement properties. Name the statistical package used and the version. | See Methods Section  See Protocol paper for Hypothesised Measurement properties:  Williams EH, Thompson NM, McCray G, Crespo-Llado MM, Bhavnani S, Gajria D, et al. Scalable Transdiagnostic Early Assessment of Mental Health (STREAM): a study protocol. BMJ Open. 2024 June 1;14(6):e088263. |
| GM8 – Missing data | Describe approaches for dealing with missing data. | Missingness, which was low, was handled via paired case omission. |
| GM9 – Unplanned analysis | Specify analyses that were unplanned and their rationale. | This is detailed in the discussion, rather than the methods:   *“Test-retest reliability indicated that STREAM scores were generally stable. Although ICC (2,1) (absolute agreement) was originally planned, ICC (3,1) (consistency) was used to account for systematic differences between sessions arising from learning effects on tablet-based tasks.”* |
| **Report section: General results** |  |  |
| GR1 – Participant characteristics | Provide study participants’ characteristics, specified per subgroup if applicable. | See Table 1 |
| GR2 – Sample size | Provide the total number of participants included in the study and the sample size for each analysis. | See Table 2 |
| GR3 – Missing data | Provide amount of (proportion or count) and reasons for missing data for each analysis for the PROM, and for any analyses of other outcome measurement instruments. | *“Missing data was 2% for PMA [Primary Measures Assessment], 1% for SMA [Secondary Measures Assessment], and 3% for longitudinal follow-up.”*  As data were obtained through non-representative convenience and quota sampling, and additional participants were recruited to meet the quota when individuals declined to take part in the study, any efforts to reduce bias through missing data imputation would have limited utility. |
| GR4 – Results | Describe the results corresponding to all objectives (see measurement properties specific boxes). | See Table 2 |
| **Report section: Discussion/conclusions** |  |  |
| DC1 – Measurement property evidence | Provide the main findings and if each measurement property is sufficient or insufficient and why. | See Discussion. |
| DC2 – Practical relevance | Discuss the practical relevance of the findings in terms of recommendations for (not) using the PROM. | See Discussion. |
| DC3 – Strengths and limitations | Discuss strengths and limitations of each study. For example, discuss if there were any potential biases in the study that could have impacted the results. | See Discussion. |
| DC4 – Generalizability | Discuss generalizability of the results. For example, discuss whether the results could be generalized to other populations given the sample studied. | We commented briefly on generalisability, stating that STREAM appears suitable for use in low-resource settings based on data from community and enriched samples in India and Malawi. However, we cannot generalise to populations or settings not represented in our sample, which would need further validation work. This is consistent with testing standards. For example, APA/AREA Standards for Educational and Psychological Testing, state:  *“Validation is the joint responsibility of the test developer and the test user. The test developer is responsible for furnishing relevant evidence and a rationale in support of any test score interpretations for specified uses intended by the developer. The test user is ultimately responsible for evaluating the evidence in the particular setting in which the test is to be used.”*  *Eignor, D. R. (2013). The standards for educational and psychological testing.* |
| DC5 – Instrument changes | Discuss what modifications are needed to the existing PROM. | Not applicable |
| DC6 – Future research | Describe new research questions or hypotheses generated from these findings, and provide/describe the research needed to answer those questions. | See Discussion. |
| **DC7 – Conclusions** | Provide the overall conclusions for the use of the PROM. | See Discussion. |
| **Report section: Other information** |  |  |
| O1 – Conflict of interest | State any conflict of interest you may have related to the PROM. This may include any involvement in the development of the PROM or any commercial funding or profit. | This will be submitted as part of the information gathered by the journal from all coauthors. |

### Supplementary Materials 2 – Recruitment Strategy

Supplementary Table 2.1. Age- and sex-stratified recruitment strategy for the community sample

|  | **0-3 mths** | **3-6 mths** | **6-9 mths** | **9-12 mths** | **1-1·5 yrs** | **1·5-2 yrs** | **2-2·5 yrs** | **2·5-3 yrs** | **3-3·5 yrs** | **3·5-4 yrs** | **4-4·5 yrs** | **4·5-5 yrs** | **5-5·5 yrs** | **5·5-6 yrs** | **Total** |
| --- | --- | --- | --- | --- | --- | --- | --- | --- | --- | --- | --- | --- | --- | --- | --- |
| **Males** | 77 | 77 | 77 | 77 | 154 | 154 | 154 | 154 | 154 | 154 | 154 | 154 | 155 | 155 | **1850** |
| **Females** | 77 | 77 | 77 | 77 | 154 | 154 | 154 | 154 | 154 | 154 | 154 | 154 | 155 | 155 | **1850** |
| **Total** | **154** | **154** | **154** | **154** | **308** | **308** | **308** | **308** | **308** | **308** | **308** | **308** | **310** | **310** | **3700** |

** This was a general guide due to anticipated difficulties in recruiting exactly the numbers reported in each age bracket.*

Supplementary Table 2.2. Age- and sex-stratified recruitment strategy for the enriched sample.

|  | **0-2 yrs** | **2-4 yrs** | **4-6 yrs** | **Total** |
| --- | --- | --- | --- | --- |
| **Targeted NDDs** | GDD | GDD, ID, ASD | ID, ASD, ADHD |  |
| **Males** | 50 | 50 | 50 | **150** |
| **Females** | 50 | 50 | 50 | **150** |
| **Total** | **100** | **100** | **100** | **300** |

** This was a general guide due to anticipated difficulties in recruiting exactly the numbers reported in each age bracket. A diagnosis for any of these conditions is not necessary for inclusion in the study. GDD = General Developmental Delay; ID = Intellectual Difficulty; ASD = Autism Spectrum Disorder; ADHD = Attention Deficit Hyperactivity Disorder.*

### Supplementary Materials 3 – Enriched Sample Details

Supplementary Table 3 Neurodevelopmental profiles within the enriched sample

|  | **India (n)** | **Malawi (n)** |
| --- | --- | --- |
| **Autism Spectrum Disorder (ASD)** | 63 | 19 |
| **Global Developmental Delay (GDD)** | 24 | 43 |
| **Cerebral Palsy (CP)** | 8 | 40 |
| **Intellectual Difficulty (ID)** | 1 | 54 |
| **Epilepsy** | 8 | 57 |
| **Attention Deficit Hyperactivity Disorder (ADHD)** | 1 | 3 |
| **Other*** | 6 | 14 |

**Other includes conditions such as hydrocephalus, behaviour problems, speech delay, etc. N.B. Children could have more than one diagnosis.*

### Supplementary Materials 4 – STREAM-Suite Tasks and Metrics

STREAM-Suite Metric Generation

START tasks were processed according to their original guidelines,^1^ which generated multiple metrics per task. These metrics were then combined to form STREAM-suite metrics via iterative factor analyses and input from Subject Matter and Content Experts to minimise dimensionality (i.e. to uncover robust and unique factors which clearly measured specific constructs). STREAM suite metrics for the MDAT and DEEP tools were generated via Item Response Theory Modelling.^2^ For MDAT, scores were modelled for each domain and overall (i.e., data from all four domains). For DEEP, we derived a single score based on i) highest level reached, ii) accuracy, and iii) completion time (see ^3^ for details). Both MDAT and DEEP were scored via a two-parameter logistic model fitted using full information factor analysis in the R-Package MIRT.^4^

1. Dubey I, Bishain R, Dasgupta J, Bhavnani S, Belmonte MK, Gliga T, Mukherjee D, Lockwood Estrin G, Johnson MH, Chandran S, Patel V. Using mobile health technology to assess childhood autism in low-resource community settings in India: An innovation to address the detection gap. Autism. 2024 Mar; 28(3):755-69.
2. De Ayala RJ. The theory and practice of item response theory. Guilford Publications; 2013 Oct 15.
3. Bhavnani S, Ranjan A, Mukherjee D, Divan G, Prakash A, Yadav A, Lal C, Gajria D, Irfan H, Sharma KK, Todkar S. A non-specialist worker delivered digital assessment of cognitive development (DEEP) in young children: a longitudinal validation study in rural India. PLOS Digital Health. 2025 May 16;4(5):e0000824.
4. Chalmers RP. mirt: A multidimensional item response theory package for the R environment. Journal of statistical Software. 2012 May 24; 48:1-29.

Supplementary Table 4.1. Full list of tasks included in the STREAM platform

| **Task** | **Ages** | **Construct** | **Administration Mode** |
| --- | --- | --- | --- |
| MDAT - Social grid | 0-6 | Social and communication | Caregiver report, observation, child assessment |
| MDAT - Language grid | 0-6 | Attention and cognition | Caregiver report, observation, child assessment |
| MDAT - Fine Motor grid | 0-6 | Motor | Caregiver report, observation, child assessment |
| MDAT - Gross Motor grid | 0-6 | Motor | Caregiver report, observation, child assessment |
| START – Preferential Looking Task | 0-6 | Social and communication | Child assessment |
| START – Parent Child Interaction | 0-6 | Social and communication | Child assessment |
| START - Button Task | 2·5-6 | Social and communication | Child assessment |
| START - Bubble Popping Task | 2·5-6 | Attention and cognition | Child assessment |
| START - Colouring Task | 2·5-6 | Motor | Child assessment |
| START - Motor Following Task | 2·5-6 | Motor | Child assessment |
| START - Synchrony Task | 2·5-6 | Motor | Child assessment |
| START - Delayed Gratification Task | 2·5-6 | Attention and cognition | Child assessment |
| START - Language Sampling Task | 2·5-6 | Attention and cognition | Child assessment |
| START - Wheel Task | 2·5-6 | Attention and cognition | Child assessment |
| DEEP - Single Tap | 2·5-6 | Attention and cognition | Child assessment |
| DEEP - Alternate Tap | 2·5-6 | Attention and cognition | Child assessment |
| DEEP - Popping Bubbles | 2·5-6 | Attention and cognition | Child assessment |
| DEEP - Grow your Garden | 2·5-6 | Attention and cognition | Child assessment |
| DEEP - Hidden Objects | 2·5-6 | Attention and cognition | Child assessment |
| DEEP - Odd one Out | 2·5-6 | Attention and cognition | Child assessment |
| DEEP - Spot the Difference | 2·5-6 | Attention and cognition | Child assessment |
| DEEP - Matching Shapes | 2·5-6 | Attention and cognition | Child assessment |
| DEEP - Jigsaw | 2·5-6 | Attention and cognition | Child assessment |
| DEEP - Sorting Objects | 2·5-6 | Attention and cognition | Child assessment |
| DEEP - Series Completion | 2·5-6 | Attention and cognition | Child assessment |
| DEEP - Pattern Making | 2·5-6 | Attention and cognition | Child assessment |
| DEEP - Sequence Recall | 2·5-6 | Attention and cognition | Child assessment |

Supplementary Table 4.2. Full list of STREAM task metrics and whether each was retained or excluded in the final model.

| **Task Name** | **Description of Task** | **Derived Metrics** | **Metrics Included in SEM** |
| --- | --- | --- | --- |
| MDAT: Gross Motor | Observation and parent report checklist assessing child's ability to complete different GROSS MOTOR related tasks (39 items).    The starting item is determined by the child's age. The grid is complete once there are 6 contiguous fails at the higher end (or the end of the grid is reached), and 6 contiguous passes at the lower end (or the start of the grid is reached). | Each attempted item is coded as either YES or NO (i.e. pass or fail).  A total score is created for the grid based on the pass rate. | GM grid total score |
| MDAT: Fine Motor | Observation and parent report checklist assessing child's ability to complete different FINE MOTOR related tasks (42 items).  The starting item is determined by the child's age. The grid is complete once there are 6 contiguous fails at the higher end (or the end of the grid is reached), and 6 contiguous passes at the lower end (or the start of the grid is reached). | Each attempted item is coded as either YES or NO (i.e. pass or fail).  A total score is created for the grid based on the pass rate. | FM grid total score |
| MDAT: Language | Observation and parent report checklist assessing child's ability to complete different LANGUAGE related tasks (40 items).  The starting item is determined by the child's age. The grid is complete once there are 6 contiguous fails at the higher end (or the end of the grid is reached), and 6 contiguous passes at the lower end (or the start of the grid is reached). | Each attempted item is coded as either YES or NO (i.e. pass or fail).  A total score is created for the grid based on the pass rate. | L grid total score |
| MDAT: Social | Observation and parent report checklist assessing child's ability to complete different SOCIAL related tasks (36 items).  The starting item is determined by the child's age. The grid is complete once there are 6 contiguous fails at the higher end (or the end of the grid is reached), and 6 contiguous passes at the lower end (or the start of the grid is reached). | Each attempted item is coded as either YES or NO (i.e. pass or fail).  A total score is created for the grid based on the pass rate. | S grid total score |
| DEEP: Single Tap | A single balloon is presented on the screen; the ballon keeps reappearing when popped until the game is complete.  Primary instructions: ‘Look what happens when I touch this balloon to pop it. Wow, look at how these stars are coming out. Now you pop the balloon. Keep popping, don’t stop popping, pop as fast as you can.’ | Accuracy (correct taps proportional to total taps);  Completion time (total time to complete game);  Latency (delay between onset and first tap). | A unidimensional DEEP total score was calculated across tasks based on accuracy, completion time, and highest level reached. |
| DEEP: Alternate Tap | Two balloons are presented side-by-side on screen. Each balloon fades into a shadow when tapped while the alternate balloon retains/regains its colour.  Primary instructions: ‘look what happens when I pop this balloon, it has gone here, now look when I pop it here (right balloon), it has gone here (left balloon). And now, when I pop it here (left balloon), it goes here (right balloon). Now you try.’ | Accuracy (correct taps proportional to total taps);  Completion time (total time to complete game);  Latency (delay between onset and first tap). | A unidimensional DEEP total score was calculated across tasks based on accuracy, completion time, and highest level reached. |
| DEEP: Popping Balloons | Balloons spawn from the bottom of the screen and drift upwards. Balloons pop when tapped. Levels (2): increased speed of spawning balloons.  Primary instructions: ‘Come let’s burst all these balloons, I will pop as many as I can. Now you try.’ | Accuracy (correct taps proportional to total taps);  Completion time (total time to complete game);  Latency (delay between onset and first tap). | A unidimensional DEEP total score was calculated across tasks based on accuracy, completion time, and highest level reached. |
| DEEP: Grow Your Garden | Apples and Owls appear randomly on the screen; when tapped, they result in a glass of juice either appearing or disappearing, respectively. Levels (4): Stimuli presentation changes from distinct to overlapping and presentation duration reduces from 3 seconds to 1 second.  Primary instructions: ‘In this game, you need to fill this place with glasses of juice. Look, when I touch an apple, a glass of juice comes, when I touch a bird, the glass of juice disappears. We have to fill this space with glasses of juice by touching the apples and not the bird. Look, I’m going to fill this space with glasses of juice by touching an apple and, another apple, and another. I’m not going to touch a bird at all, just the apple so that I can fill this space with glasses of juice. Now you try and fill up this space with glasses of juice.’ | Accuracy (correct taps proportional to total taps);  Completion time (total time to complete game);  Latency (delay between onset and first tap). | A unidimensional DEEP total score was calculated across tasks based on accuracy, completion time, and highest level reached. |
| DEEP: Hidden Objects | Multiple animals simultaneously hide in different locations. The child must touch the correct hiding places. Levels (5): Increasing number of hiding characters.  Primary instructions: ‘Look at these 2 birds carefully, they are going to hide in these trees. Let’s remember where have they hidden. Look at where they have hidden, I will touch the trees to take them out, here it is. Now I’ll find the second one, here it is. Now you try. | Accuracy (correct taps proportional to total taps);  Completion time (total time to complete game);  Latency (delay between onset and first tap). | A unidimensional DEEP total score was calculated across tasks based on accuracy, completion time, and highest level reached. |
| DEEP: Odd One Out | Four objects are presented; three are identical and one is distinct. The child must touch the distinct object. Levels (4): Objects differed on either colour, size, shape or category.  Primary instructions: ‘Look at these 4 objects carefully. Out of these 4 objects, 3 are similar and 1 is different. I am only going to tap the one which is different from the others. Now you try.’ | Accuracy (correct taps proportional to total taps);  Completion time (total time to complete game);  Latency (delay between onset and first tap). | A unidimensional DEEP total score was calculated across tasks based on accuracy, completion time, and highest level reached. |
| DEEP: Spot the Difference | Two images are presented on the screen. The child must touch the missing objects to make the two images identical.  Primary instructions: ‘Look at these boxes. See these objects in the boxes. There are many objects here and only 1 object here. We have to touch those objects which are here (left box) but not here (right box). When I touch those objects, they also come here (assessor to touch objects in the left box and show that they appear in the right box). We have to make these 2 boxes identical. Now you try.’ | Accuracy (correct taps proportional to total taps);  Completion time (total time to complete game);  Latency (delay between onset and first tap). | A unidimensional DEEP total score was calculated across tasks based on accuracy, completion time, and highest level reached. |
| DEEP: Matching Shapes | Objects are presented on the left with their outlines on the right. The child has to drag and drop the objects to their outline. Levels (5): Increasing similarity between objects presented together.  Primary instructions: ‘Look at these 2 objects carefully. Look at this vest, this is the vest shadow, just like this vest. Put this vest on its shadow. Touch this, drag it and leave it on its shadow. Look at these shorts, this is the shadow of these shorts, put these shorts on their shadow. Now you try, put all the objects on their shadow.’ | Accuracy (correct taps proportional to total taps);  Completion time (total time to complete game);  Latency (delay between onset and first tap). | A unidimensional DEEP total score was calculated across tasks based on accuracy, completion time, and highest level reached. |
| DEEP: Jigsaw | Animal parts are presented on the left with the outline of the complete animal on the right. Each part has to be moved to its outline to create the whole animal. Levels (6): Increased number of pieces of jigsaw and similarity between them.  Primary instructions: ‘Look at this carefully, it is the face of the giraffe. Like we did earlier, let’s put the face of the giraffe here, onto the face. Let’s put the tail of the giraffe here, onto the tail. Now you try.’ | Accuracy (correct taps proportional to total taps);  Completion time (total time to complete game);  Latency (delay between onset and first tap). | A unidimensional DEEP total score was calculated across tasks based on accuracy, completion time, and highest level reached. |
| DEEP: Sorting Objects | Objects and containers with labels relevant to different objects are presented on the screen. The child has to drag and drop the objects into the correct container.  Primary instructions: 'Look at these objects carefully. Take them where they belong according to their shape. See, this one is a square, so it will go here, this one is a circle so it will go here. Now I will touch, drag and drop the star here, and I will touch, drag and drop the circle here. Now I will take each of these objects to their location. Now you try.’ | Accuracy (correct taps proportional to total taps);  Completion time (total time to complete game);  Latency (delay between onset and first tap). | A unidimensional DEEP total score was calculated across tasks based on accuracy, completion time, and highest level reached. |
| DEEP: Series Completion | An incomplete sequence of objects is presented alongside different objects which can be selected. Child has to drag and drop the correct object to complete the sequence.  Primary instructions: ‘Look at these objects. There is a blue object, then a red object, after the red comes blue, then comes red again, what will come next? Blue and then? Red. I will take the blue object from here (point to a row of objects at the bottom), touch it, drag it and drop it here. What will come next? Red. I will take the red object from here (point to a row of objects at the bottom), touch it, drag it and drop it here. And I will complete this line. Now you try.’ | Accuracy (correct taps proportional to total taps);  Completion time (total time to complete game);  Latency (delay between onset and first tap). | A unidimensional DEEP total score was calculated across tasks based on accuracy, completion time, and highest level reached. |
| DEEP: Pattern Making | The child has to copy the pattern by touching, dragging and dropping square pieces to make the big box identical to the reference box.  Primary instructions: ‘Look at this box carefully. The first row has a red line, the second row has a green line and the third row has a blue line. I have to make the same type of box here. I will touch, drag and drop the red colour square on this, this and this and complete this line. Like this, I will touch, drag and drop the green colour on this, this and this and complete this line. Then I will touch, drag and drop the blue colour on this, this and this and complete this line. Like this, I will make this box exactly like that one. Now you try.’ | Accuracy (correct taps proportional to total taps);  Completion time (total time to complete game);  Latency (delay between onset and first tap). | A unidimensional DEEP total score was calculated across tasks based on accuracy, completion time, and highest level reached. |
| DEEP: Sequence Recall | Squares in a grid light up sequentially, which need to then be tapped by the child in the same order.  Primary instructions: ‘Look at these boxes carefully. See how they light up. First, this one lights up, then this one. The one which lights up first, I will press that first, the one which lights up second, I will press that next. Now you try.’ | Accuracy (correct taps proportional to total taps);  Completion time (total time to complete game);  Latency (delay between onset and first tap). | A unidimensional DEEP total score was calculated across tasks based on accuracy, completion time, and highest level reached. |
| DEEP: Location Recall | This game 'bookends' each of the games described above. Before each game listed above, the moon is shown moving to hide behind one of multiple hiding places. The child has to recall the moon’s hiding location after completing each game.  Primary instructions: 'See here, this moon is hiding from [CHILD’S NAME], it has hidden, where has it hidden, I will touch that place and get the moon out. See where the moon is hiding. Now you touch the place and get the moon out.' | Accuracy (correct taps proportional to total taps);  Completion time (total time to complete game);  Latency (delay between onset and first tap). | A unidimensional DEEP total score was calculated across tasks based on accuracy, completion time, and highest level reached. |
| START: Wheel Task | A video of a black and white spinning wheel is displayed on the screen across multiple trials. The child is able to tap a button to stop the video, which will skip to the next trial. Thus, if the child enjoys the spinning wheel stimulus and wants to watch for longer, they can choose to NOT press the button.  Primary instructions: "Look at this wheel moving. You can look at it for as long as you want. If you want to stop it just press this red button." | Total time spent looking at the wheel;  Mean distance of the child's face from the screen;  SD of distance of the child's face from the screen. | Removed due to low factor loading (<0.34)  Removed due to low factor loading (<0.34)  Removed due to low factor loading (<0.34) |
| START: Preferential Looking Task | Across several trials, a brief social video is displayed on one half of the screen while a brief non-social video is displayed on the other half of the screen (presentation side is counterbalanced across stimulus type).  Primary instructions: “I would like to show you something on my tablet. Look at these videos. Try to stay still and keep looking at the screen.” | Proportion of gaze frames directed at social stimuli out of all gaze frames on either stimulus. | Removed due to low factor loading (<0.34) |
| START: Button Task | On each trial the child can choose to tap either a green button or a red button: one button will always show a social video when pressed; the other button will always show a non-social video when pressed.  Primary instructions: “Look at this…if we press this red button, a (describe video) appears. And if we press this green button, a (describe video) appears. I want to watch a child (press the button that contains a video of a child). Now I want to watch an 'object' (press the button that contains the video of moving lines). Which button do you think has the child? Which button do you think has the 'object'?” | Ratio of the number of trials that the child chose the social relative to the non-social video. | Removed due to low factor loading (<0.34) |
| START: Colouring Task | An uncoloured picture is displayed. The child can choose from different colours to colour in the image using their finger.  "Look at this picture. Look at all these colours I can choose to colour it in.” After colouring the image partially, ask the child to colour in the same way. “Can you take a colour and colour in this picture without leaving the lines?" | Colouring quality score: A measure of drawing accuracy, calculated as the proportion of coloured pixels inside the target area adjusted for the proportion coloured outside. Higher scores indicate that the child coloured more within the target and less outside. | Retained |
| START: Motor Following Task | On each trial, a butterfly starts on one side of the screen and travels to the other side following a different 'sine wave-esque' trajectory, with occasional fluctuations in speed and direction. The child must touch the screen and follow the path of the butterfly using their finger.  Primary instructions: "See how I am trying to chase the butterfly? It goes up…it goes down…Look I caught it!" (tapping on the butterfly at the end). Keep the child’s attention on the screen. After the demo is finished, say to the child "Can you chase the butterfly with your finger?" | RMSE: Spatio-temporal difference between the target and the child’s motor trajectory;  Weighted_x_freq_gain: Frequency gain x to assess the closeness in the source and target motions along the frequency domain;  Weighted_y_freq_gain: Frequency gain y  Speed: Average rate of change in touch position over time, reflecting how fast the child's finger moved across screen  Acceleration: Average rate of change in speed over time, reflecting how the child increased or decreased speed;  Jerk: Average rate of change in acceleration over time, reflecting the smoothness or abruptness of finger movements. | Retained  Retained  Retained  Retained  Retained  Retained |
| START: Bubble Popping Task | Bubbles appear in the screen and move up and down; the child must pop the bubbles by tapping them with their finger.  Primary instructions: "See I am going to touch these bubbles to pop them, here they go pop, pop, pop.” After popping a few bubbles, say to the child: “Now you play." | Completion time (how long it takes the child to pop all bubbles);  Tap precision (distance of child's finger tap from the centre of the bubble). | Retained  Removed due to low factor loading (<0.34) |
| START: Synchrony Task | A child avatar hitting a drum at a steady rhythm is displayed on screen. The child must tap their drum, shown at the bottom of the screen, in time with the avatar's drum taps. The avatar changes to a different steady rhythm part way through.  Primary instructions: “Look at this child. He wants to drum with you. Listen to how he drums. Try to tap your drum at the same time as this child.” | Mean resultant length: a 0-1 index of how tightly the child’s tap timings cluster around the average phase (higher score = better synchrony). | Removed due to low factor loading (<0.34) |
| START: Delayed Gratification Task | The child is shown a screen with a 'loading wheel' in the centre and a gold star in the corner. The child can choose to tap the star at any point to receive 1 reward immediately, or they can wait for 3 minutes (during which nothing of interest happens on the screen) and receive 3 rewards. The child does not know how long they will have to wait; the only clue is a 'loading wheel' on the screen which loads for a short while but then seemingly becomes 'stuck' and remains that way until the 3 minutes is over.  Primary instructions: "Look at these! (show the reward) You can have 3 of these if you wait until this finishes (point to the wheel icon). If you don't want to wait, press the yellow star (point to yellow star) but then you only get 1." | Duration of time that child waited (i.e. time from onset of task to the star being tapped, up to a maximum of 3 mins if the child did not tap the star). Corrected to account for instances where the child was not sat in front of the screen (duration waited * proportion of frames in which child’s face was detected). | Removed due to low factor loading (<0.34) |
| START: Language Sampling Task | Child is shown an image in which lots of different things are depicted. The child is asked to describe the different things they can see in the image.  Primary instructions: 'Look at this picture. There are lots of different things happening in the picture’ “Can you tell me about what's happening in this picture?" | Analysis plan not yet confirmed. | This task was not included in the SEM as manual coding could not be completed in time. |
| START: Parent-Child Interaction | Parent and child are given a set of toys and are asked to play as they would at home. After around 5 minutes of 'freeplay' the parent disengages from the child for 2 minutes to see how the child reacts to this unusual situation.  Primary instructions: “For the next 5 minutes, we would like to watch you and your child interacting. Here are some toys you can play with [show the mother the toys]. You can play with as many or as few of the toys as you like. If [child’s name] shows a preference for one of the toys and wants to play with it for the entire interaction, that is ok. If your child is not interested in any of the toys, you can interact together without toys.  "After 5 minutes of interaction, I will clap my hands. For the next 2 minutes, I would like you to look away from your child and ignore him/her. It is important that you keep an upset face and both of your arms crossed on your chest. If while ignoring your child, he/she approaches you, please respond to your child as briefly as possible and then continue to ignore him/her. It is essential that during those 2 minutes you are able to disengage from your child as much as possible." | A global coding scheme was adopted, wherein local raters at each site scored the constructs noted below using a 5-point Likert scale (1 - Never, or only for a very brief period; 2 = Rarely; 3 = Sometimes; 4 = Often; 5 = All the time, or nearly all the time).  *Attentiveness to Caregiver*: The infant's level of interest in their caregiver as demonstrated by the amount of time they look toward their caregiver (either face or body), listen and/or respond to their caregiver, and their acceptance of and engagement with their caregiver's involvement in the interaction.  *Effective Communication*: The quantity/quality of the infant's communicative action directed toward the caregiver. Communicative actions could be verbal, but may also include movement of limbs, gestures (e.g. pointing with a digit), non-speech vocalisations, and facial expressions directed to the caregiver. Crying and 'maladaptive' communicative behaviours (e.g. aggressive behaviours toward caregiver, such as pulling hair) would not be coded as effective communication.  *Liveliness*: The level of physical activity displayed by the child. Activity does not necessarily mean the child is standing and/or moving around; rating includes bodily/limb movements while seated, engagement with toys. Liveliness primarily considers the frequency of physical activity, but some consideration is also given to the intensity of activity.  *Positive Affect*: The child's positive affect, ranging from a child who never displays any positive emotion to one who continually exhibits signs of positive emotion (i.e. joy, excitement, happiness). This includes positive vocalisations, gestures displaying excitement (e.g., clapping, raising hands), smiles, laughter, and happy facial expressions.  *Negative Affect*: The child's negative affect, ranging from a child who never displays any negative emotion, to one who continually exhibits signs of negative emotion (i.e. distress, sadness, frustration, anger). This includes crying, screaming, kicking, and angry/sad facial expressions.  *Infant-Caregiver Engagement*: The quantity/quality of dyadic engagement during the session. Dyadic engagement includes episodes where the mother and child are interacting and/or focusing on the same object/toy by looking at or otherwise engaging with it (i.e. joint attention episodes), looking at each other (i.e. mutual gaze episodes), displaying matching emotions during play (e.g. exchange of smiles, shared enjoyment), and/or are engaging in turn-taking episodes (e.g. mother singing/talking and child responding back or vice versa). Unlike the other codes, this code considers the behaviour of the child AND the caregiver to the extent that they both affect the engagement/interaction. | This task was used as an exploratory convergent measure and was not included in the derivation of the STREAM scores. |

### Supplementary Materials 5 - Modelling

Supplementary Table 5. Model fit statistics.

|  | **Model 1** | **Model 2** | **Model 4** |
| --- | --- | --- | --- |
| **Chisq (P-Value)** | 65·02 (16) - p<0·001 | 58·63 (13) - p<0·001 | 63·56 (15) - p<0·001 |
| **RMSEA (95%CIs)** | 0·029 (0·022-0·036) | 0·031 (0·023-0·039) | 0·029 (0·022-0·037) |
| **CFI** | 0·986 | 0·987 | 0·987 |
| **TLI** | 0·976 | 0·973 | 0·975 |
| **Akaike (AIC)** | 52471·961 | 52471·566 | 52472·497 |

*Note: Models 3 and 5 are not presented as there were no theoretically justified cross-loadings based on modification indices.*

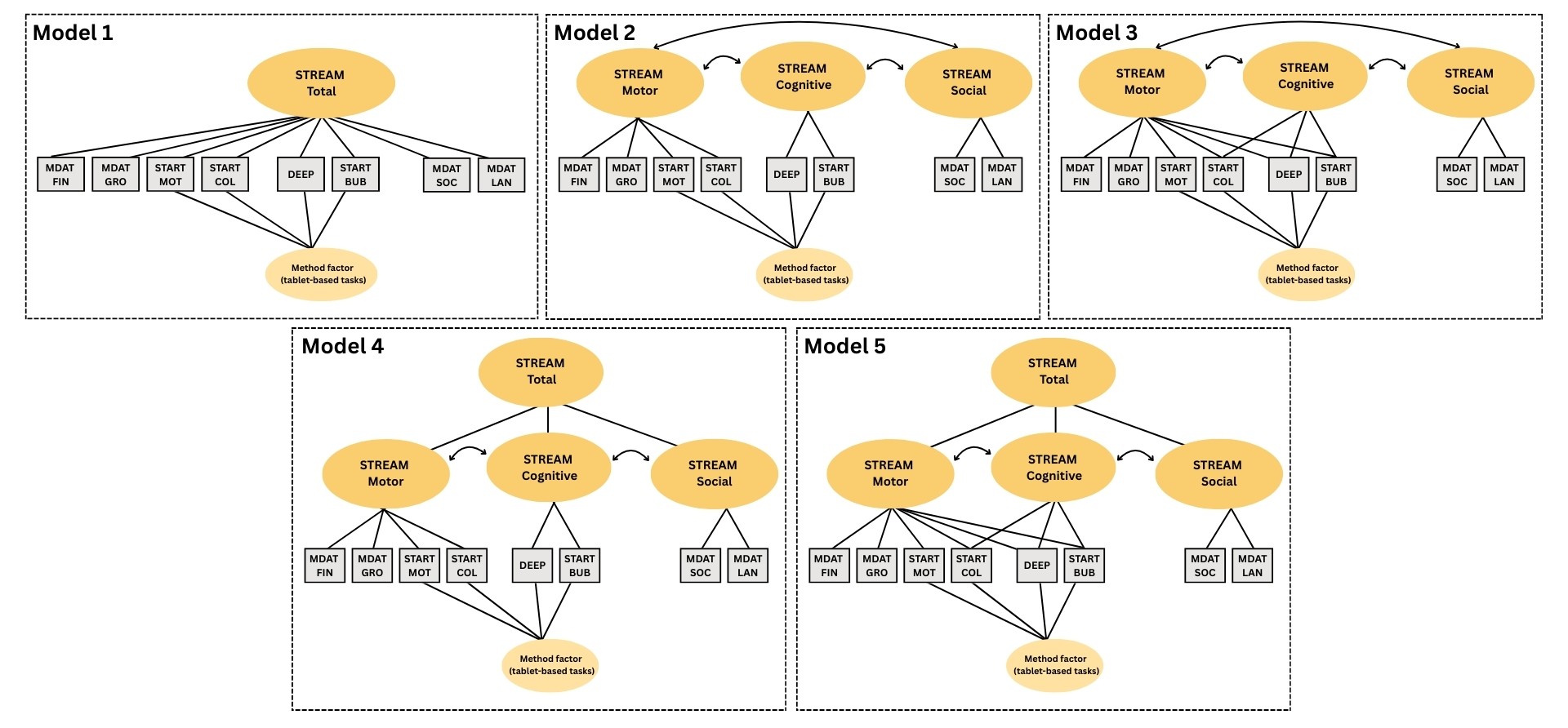
Supplementary Figure 5. Illustration of competing models tested during model fitting.

Following our published protocol (Williams et al., 2024), several competing models were tested (Supplementary Figure 5). Model 1, the baseline comparator, was unidimensional with all items loading on one factor. Model 2 ascribed one task to one domain (i.e., no cross-loading). Model 3 allowed cross loadings. Model 4 did not allow cross loadings and included an additional second-order factor representing a ‘total score’. Model 5 allowed cross loadings and included an additional second-order factor representing a ‘total score’.

### Supplementary Materials 6 – Additional Psychometric Properties

Supplementary Table 6. Additional psychometric properties

| **Evidence type** | **Construct** | **Measure** | **Statistic** | **Sample size** | **Ind** | **Mal** | **India** | **Malawi** | **India** | **Malawi** | **India** | **Malawi** |
| --- | --- | --- | --- | --- | --- | --- | --- | --- | --- | --- | --- | --- |
|  |  |  |  |  | **Achieved N** | | **Motor** | | **Attention & Cognition** | | **Social & Communication** | |
| **Convergent validity** | Parent-child relationship | MORS (0-3 years) (invasion) | Pearson | 2000 | 930 | 916 | -0·06  (-0·13-0·01) | -0·04  (-0·10-0·02) | -0·07  (-0·13-0·00) | -0·04  (-0·10-0·03) | -0·06  (-0·12-0·01) | -0·04  (-0·10-0·02) |
|  |  | CPRS (3-6 years) (conflict) | Pearson | 2000 | 924 | 852 | **-0·09**  **(-0·15--0·03)** | **-0·17**  **(-0·23--0·10)** | **-0·09**  **(-0·15--0·03)** | **-0·16**  **(-0·23--0·10)** | **-0·09**  **(-0·16--0·03)** | **-0·17**  **(-0·23--0·11)** |
|  |  | Attention to caregiver (freeplay) | Pearson | NA | 287 | 270 | **0·15**  **(0·03-0·27)** | -0·03  (-0·16-0·09) | **0·16**  **(0·02-0·27)** | -0·03  (-0·15-0·09) | **0·15**  **(0·04-0·26)** | -0·03  (-0·15-0·09) |
|  |  | Communication (freeplay) | Pearson | NA | 286 | 270 | **0·16**  **(0·05-0·28)** | 0·02  (-0·10-0·14) | **0·16**  **(0·04-0·27)** | -0·01  (-0·14-0·12) | **0·17**  **(0·05-0·27)** | 0·02  (-0·1-0·15) |
|  |  | Liveliness (freeplay) | Pearson | NA | 287 | 270 | 0·08  (-0·03-0·19) | 0·07  (-0·04-0·21) | 0·10  (-0·03-0·22) | 0·05  (-0·08-0·17) | 0·09  (-0·04-0·21) | 0·08  (-0·04-0·19) |
|  |  | Positive Affect (freeplay) | Pearson | NA | 287 | 270 | **0·26**  **(0·15-0·37)** | -0·07  (-0·19-0·05) | **0·27**  **(0·15-0·38)** | -0·09  (-0·22-0·04) | **0·27**  **(0·16-0·38)** | -0·07  (-0·19-0·06) |
|  |  | Negative Affect (freeplay) | Pearson | NA | 287 | 270 | **-0·15**  **(-0·26--0·03)** | -0·07  (-0·20-0·06) | -0·12  (-0·25-0·01) | -0·06  (-0·19-0·08) | **-0·15**  **(-0·27--0·02)** | -0·07  (-0·19-0·06) |
|  |  | Interaction (freeplay) | Pearson | NA | 287 | 270 | **0·24**  **(0·13-0·34)** | -0·01  (-0·14-0·11) | **0·24**  **(0·12-0·35)** | -0·04  (-0·16-0·09) | **0·23**  **(0·12-0·35)** | -0·01  (-0·13-0·10) |
|  |  | Attention to caregiver (disengagement) | Pearson | NA | 286 | 270 | 0·1  (-0·01-0·21) | -0·02  (-0·13-0·10) | 0·11  (0·00-0·22) | -0·01  (-0·13-0·12) | 0·11  (0·00-0·22) | -0·02  (-0·14-0·10) |
|  |  | Communication (disengagement) | Pearson | NA | 286 | 270 | 0·03  (-0·08-0·14) | 0·03  (-0·10-0·15) | 0·04  (-0·08-0·16) | 0·03  (-0·08-0·15) | 0·04  (-0·08-0·15) | 0·03  (-0·09-0·14) |
|  |  | Liveliness (disengagement) | Pearson | NA | 286 | 270 | 0·07  (-0·05-0·19) | -0·03  (-0·16-0·10) | 0·09  (-0·03-0·21) | -0·05  (-0·17-0·06) | 0·07  (-0·05-0·19) | -0·04  (-0·16-0·08) |
|  |  | Positive Affect (disengagement) | Pearson | NA | 286 | 270 | **0·11**  **(0·00-0·22)** | -0·03  (-0·15-0·09) | **0·13**  **(0·01-0·23)** | -0·04  (-0·17-0·07) | **0·11**  **(-0·01-0·23)** | -0·03  (-0·15-0·09) |
|  |  | Negative Affect (disengagement) | Pearson | NA | 285 | 270 | -0·08  (-0·19-0·03) | 0·00  (-0·15-0·14) | -0·09  (-0·20-0·03) | -0·03  (-0·18-0·11) | -0·08  (-0·19-0·04) | -0·01  (-0·16-0·14) |

### Supplementary Materials 7 – Acknowledgements

We are grateful to the field teams who supported data collection. In Malawi, we thank Gracium Kasawala, Petros Gladwell Munthali, Priscilla Kamangira, Chikumbutso Kayimba, Grace Mlenga, Mary Ali, Asante Kadama, Tawonga Mzumara Foti, Patricia Tepetheya, and Ken Chombo. In India, we thank Bipasha Bhattacharyya (Research Coordinator), Keshav Shakya (Field Officer), Renu Tyagi and Shahin Parveen (Recruitment Officers), Saleha Khatoon, Rinku Chauhan, Monika Kumari, Diksha Singh, Rakhi Sharma, and Minakshi Rawat (Assessors), Mona Kumari, Anchal Singh, and Sunita Kumari (Site Office Assistants), and Jitender Kumar Singh (Site Coordinator). We thank Praveen Maurya and Akshat Gautam for their technical input. We also thank all the participating families.

This work was supported by the Medical Research Council’s (MRC) Global Challenges Research Fund (GCRF), project reference MR/S036423/1 (PI: BC). BC acknowledges further support from the European Research Council (grant ref: 865568) and the University of Reading. MKB was supported by the Nottingham Trent University International Partnership Fund. EJ acknowledges further support from the Medical Research Council (MR/K021389/1; MR/T003057/1), and the EU-AIMS and AIMS-2-TRIALS programmes funded by the Innovative Medicines Initiative (IMI) Joint Undertaking Grant Nos. 115300 and No. 777394; European Union’s FP7 and Horizon 2020, respectively). This Joint Undertaking receives support from the European Union's Horizon 2020 research and innovation programme, with in-kind contributions from the European Federation of Pharmaceutical Industries and Associations (EFPIA) companies and funding from Autism Speaks, Autistica and SFARI. Any views expressed are those of the author(s) and not necessarily those of the funders.
